# Development of a scoring model for the Sharp/van der Heijde score using convolutional neural networks and its clinical application in predicting radiographic progression using a graph convolutional network

**DOI:** 10.1101/2022.06.08.22276135

**Authors:** Suguru Honda, Koichiro Yano, Eiichi Tanaka, Katsunori Ikari, Masayoshi Harigai

## Abstract

**Objective:** To construct a predictive model for the Sharp/van der Heijde score (SHS) and assess its applicability in clinical research settings.

**Methods:** First, we built a rheumatoid arthritis (RA) image database linked to clinical information. We then constructed the prediction model in three steps: orientation, detection, and damage prediction. We assessed whether the SHS generated by our model replicated known findings on the association between radiographic progression and serological markers. Finally, after characterizing the SHSs of 4,264 patients using hierarchical clustering, we constructed a predictive model for joint destruction using a graph convolutional network (GCN).

**Results:** We built a model with an accuracy of 100% in the correction of image orientation using EfficientNet and constructed a model with all predicted joint coordinates within 10 pixels of the correct coordinates using U-Net. In the damage prediction phase, the EfficientNet-based model combined with the modules achieved correlation coefficients of 0.879 for erosion and 0.868 for joint space narrowing between the model and expert, exceeding that of the previous best model. Our model replicated the known finding of erosion progression’s association with serologically positive patients. The areas under the curve for predicting finger and wrist erosion in the GCN model were 0.800 and 0.748, respectively. We observed that clusters generated by hierarchical clustering ranking in the top 10 were important features in the GCN for predicting erosion.

**Conclusion:** We constructed a high-performance scoring model for SHSs applicable to clinical research. Our analysis revealed that clusters are important for predicting erosion using the GCN.

**Key messages:** *What is already known about this subject?:*

► Several deep learning models that automatically predict Sharp/van der Heijde scores (SHSs) have been reported. However, the accuracy of their joint detection and erosion prediction was insufficient, and more importantly their clinical applicability was unclear.
► Models for predicting joint destruction using clinical factors have been constructed with arbitrary factor selection by humans. No report has demonstrated the usefulness of deep learning models in predicting joint destruction using large SHS datasets.

*What does this study add?:*

► Our deep learning model showed a high performance in both joint space narrowing and erosion, replicated previous findings on association between joint destruction and serological markers, thereby demonstrating, for the first time, that deep learning models could be clinically applicable in estimating SHSs.
► We also demonstrated that a graph convolutional network (GCN) is a high performance model in predicting radiographic progression.

*How might this impact clinical practice or future developments?:*

► We believe our model will be an essential tool for future studies, such as in genome-wide association studies (GWAS) for joint destruction on a scale of thousands to millions, which is difficult to achieve with human scoring. Ultimately, data from large-scale GWAS will be integrated into the GCN to build a powerful model for precision medicine.

## INTRODUCTION

Rheumatoid arthritis (RA) is an autoimmune disease caused by chronic inflammation, which induces bone erosion and cartilage loss. The ultimate goal of RA treatment is to prevent joint destruction and protect the patient’s quality of life. Accurately assessing joint destruction is important for determining the correct course of treatment and efficacy of new drugs. Therefore, several assessment methods have been considered, and the Sharp/van der Heijde score (SHS) is currently used for as a main factor for therapeutic evaluation in clinical trials.[1] However, large-scale studies, such as genome-wide association studies (GWAS) for radiographic progression, have not been conducted, despite the large amount of radiographic data available at hospitals because the SHS is labor-intensive and requires highly skilled personnel.

Deep learning models, particularly convolutional neural networks (CNNs), offer a solution to this problem. Several CNN-based scoring systems have been reported for joint destruction in RA.[2–4] However, to the best of our knowledge, automated scoring systems applicable to clinical or research settings have not been constructed owing to the following reasons. First, the previous image dataset in RA was mostly dominated by intact joint images.[2] Collecting images from recent clinical trials yields a class-imbalanced dataset with many images of intact joints and few of progressive joints because of advances in the RA treatment. Such imbalanced datasets result in poor generalized performance.[5] Second, the model struggles to recognize joints with erosion and deformation. In radiographs of hands and feet with severe erosions and deformations, joints may disappear, and identifying joints becomes difficult using standard CNNs and object detection models. Another reason is the lack of a large image database linked to clinical data. Constructing a high-performance model is not only important but also necessary for clinical application. Given that the ultimate goal is clinical application, confirming the replication of known findings using the model is equally important. However, because no image database is linked to clinical data, no deep learning model has been confirmed to reproduce known findings.

Here, we first built a database of approximately 50,000 RA images linked to clinical information from the Institute of Rheumatology, Rheumatoid Arthritis (IORRA) cohort, which is one of the largest RA cohorts in Japan. This database includes images of progressive joint destruction from the 1990s, which mitigates class imbalance in the dataset. Then, we constructed an automated scoring model in three steps: correction of image orientation, detection of joint coordinates, and damage prediction (Figure 1). We combined various modules with the base CNN model, EfficientNet,[6] to improve its performance. To further investigate whether this improvement was clinically meaningful, we evaluated whether the SHS generated by the CNN replicated known findings regarding the association between radiographic progression and serum markers. Finally, a graph convolutional network (GCN) was used to build a predictive model of joint destruction.

**Figure 1.**
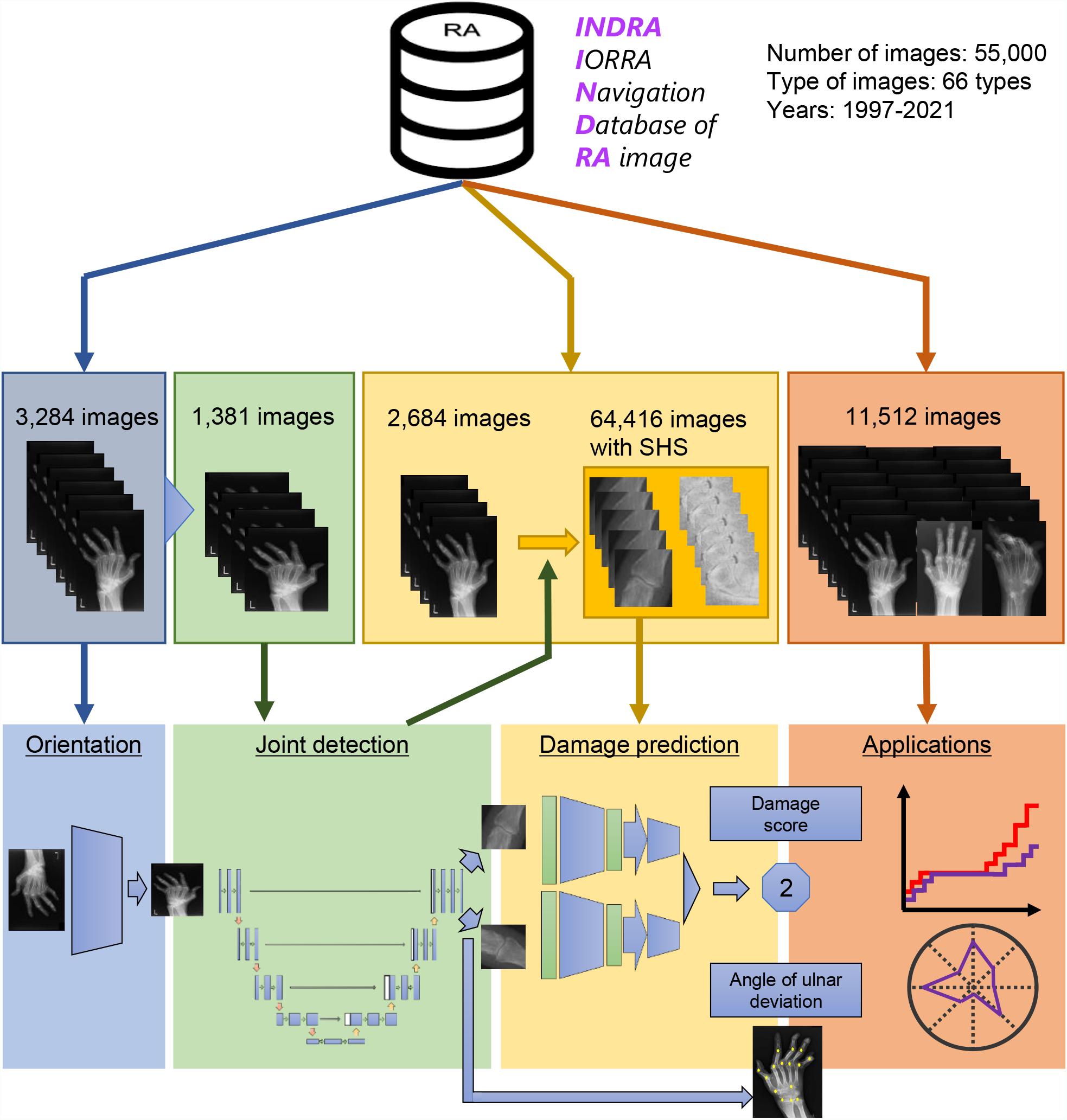
Overview of this study. The study consists of four phases: orientation, joint detection, damage prediction, and clinical and research application. All image data were extracted from INDRA. In the orientation phase, we constructed a CNN model to align image orientations. In the joint detection phase, we constructed a U-Net-based model to extract joint coordinates. The ulnar deviation angle, an indicator of rheumatoid arthritis deformity, can be calculated from the extracted joint coordinates. In the damage prediction phase, we cropped 64,416 joint images trained using an EfficientNet-based model combined with various modules. In the final phase, we first applied the model to determine whether the scores predicted by our model could replicate the known findings of the association between the radiographic progression of joint damage and serological markers. We then performed hierarchical clustering and constructed a model to predict radiographic progression using a GCN.

## METHODS

The supplementary methods provide more detail on the methods used in this study.

### Study population

All patients were enrolled in the IORRA cohort, and clinical information was obtained from this cohort. All radiographs were extracted from the IORRA Navigation Database of RA images (INDRA), which is the largest database in Japan linked to clinical data (Figure 1). For the damage prediction phase, we enrolled 1342 patients with SHSs on hand radiographs and generated 64,416 joint images. In hierarchical clustering, we extracted 4,264 individuals with at least one hand of radiographic data from INDRA. For patients with multiple radiographic data points, the oldest radiograph was extracted to use the clustering results as explanatory variables for the predictive model. In the subgroup analysis of SHSs according to serological markers and a prediction model for joint destruction using the GCN, we extracted 1503 patients with hand radiographs taken from at least two time points within INDRA.

### Convolutional neural networks in the damage prediction phase

We adopted EfficientNet as our base CNN model and combined it with various modules to improve performance (Figure 3A). To compare all models on the same test images, we first divided each joint image dataset into 10 segments, used one as the test set, and cross-validated with the remaining sets. We trained a full model EfficientNet with the following hyperparameters (optimizer: Adabound, scheduler: Cosine Annealing with T-max = 40 and learning rate from 5e-03 to 1e-06, loss function: cross entropy, epoch: 200, batch size: 32)

### Graph convolutional network in the construction of the prediction model

In our graph network, we considered each patient as a node connected by edges, based on cosine similarity. Our GCN model consists of two graph convolution layers, an inserted dropout and leaky ReLU, and a fully connected layer (Figure 7A). We trained our GCN model with the following parameters, Leaky ReLU slope of 0.1 and drop rate of 0.6.

## RESULTS

### Constructing the model to detect the orientation of hand radiographs

The first step in the automated scoring model is to detect the orientation of the hand radiographs and align them in the same direction. A total of 12,866 images, of which 9,852 were used to train EfficientNet b0,[6] and the remaining 3,284 were used to evaluate the model (Extended Figure S1A). The model achieved an accuracy of 100% in image orientation (Extended Figure S1B).

### Constructing the model to detect joint coordinates based on weakly supervised learning

We obtained 20,715 ground-truth coordinates from 1,381 hand radiographs using an in-house script (Extended Figure S2). First, we used ResNet34 [7] to directly regress the coordinates of the joints, according to a previous report. [2] However, this model could not accurately detect the joints, particularly in hands with severe joint deformities (Figure 2B). Although we changed ResNet to DenseNet [8] and EfficientNet and incorporated coordconv [9], the results did not change.

**Figure 2.**
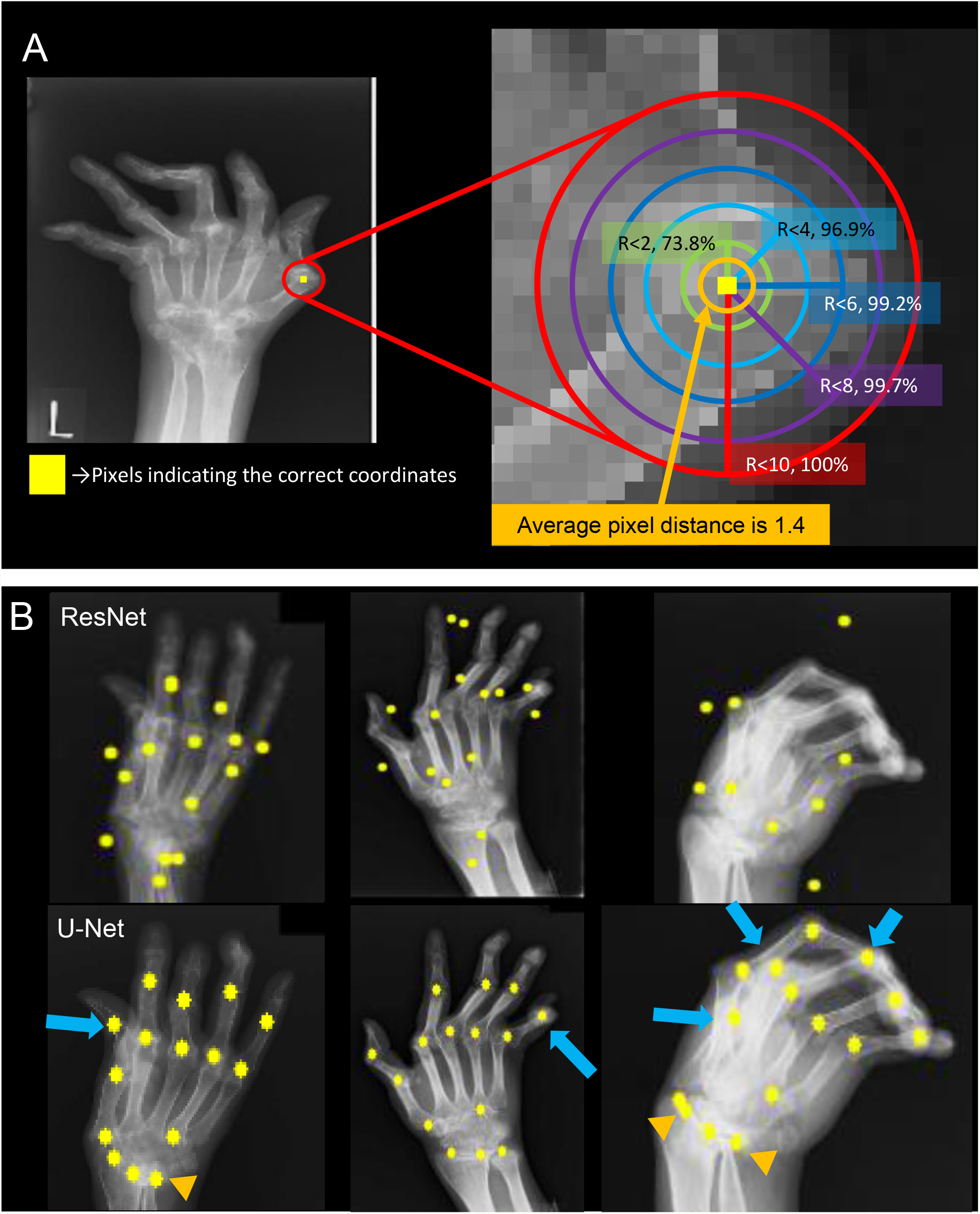
Overview of the joint detect phase using heat map regression based on U-Net. A resized radiograph with the yellow square indicating the correct coordinates of the IP joint and red circle marking a radius of 10 pixels from the coordinates (A left). An enlarged image of the area of 10 pixels from the correct coordinates is shown on the right of Panel A. The average pixel distance between the predicted and correct coordinates is 1.4 in 6215 joints (A right, orange circle). The proportion of predicted coordinates are shown contained in circles of different colors. We replotted the coordinates predicted by ResNet (upper Panel B) and U-Net (lower Panel B) on the original image. Predictions with U-Net did not deviate from the correct labels, even when the joints overlapped (B, blue arrows) or were ankylosed or collapsed (B, orange arrowhead).

Therefore, we performed weakly supervised learning by heat map regression [10–13] based on U-Net [14] (Extended Figure S3) for the joint coordinate detection. In brief, heatmap image regression does not directly regress coordinates but regresses images that encode the pseudo-probability of landmarks at a particular pixel location (Extended Figure S4 and Supplementary methods). Consequently, the average pixel distance between the 6,215 predicted and correct coordinates was 1.4 after resizing the images to 224×224 pixels. In addition, 96.9% of the predicted coordinates were within four pixels of the correct coordinates, and 100% of the predicted coordinates were within 10 (Figure 2A). We replotted the coordinates predicted by U-Net on the original hand radiographs and confirmed that U-Net accurately estimated coordinates even in the presence of severe joint deformities, overlapping joints, and joint ankylosis (Figure 2B). We cropped 64,416 joint images for the damage-prediction phase (Extended Figure S5).

### Assessment of the model in score joint destruction

We constructed our model by combining EfficientNet b3 with the following modules: over-sampling,[5] transfer learning, squeeze-and-excitation blocks,[15] self-mutual learning,[16] adabound,[17] and cosine annealing [18] (Figure 3A). Our model achieved average correlation coefficients of 0.868 for joint space narrowing (JSN) and 0.879 for erosion in the test set and root mean squared error (RMSE) of 0.664 for the JSN, 0.687 for the erosion in test-set. Although a simple comparison is not possible owing to the different methods of regression, class-balance, and dataset sizes, the correlation coefficients of our model, particularly erosion predictions, outperformed those of previously reported models (Table 1).[2]

**Figure 3.**
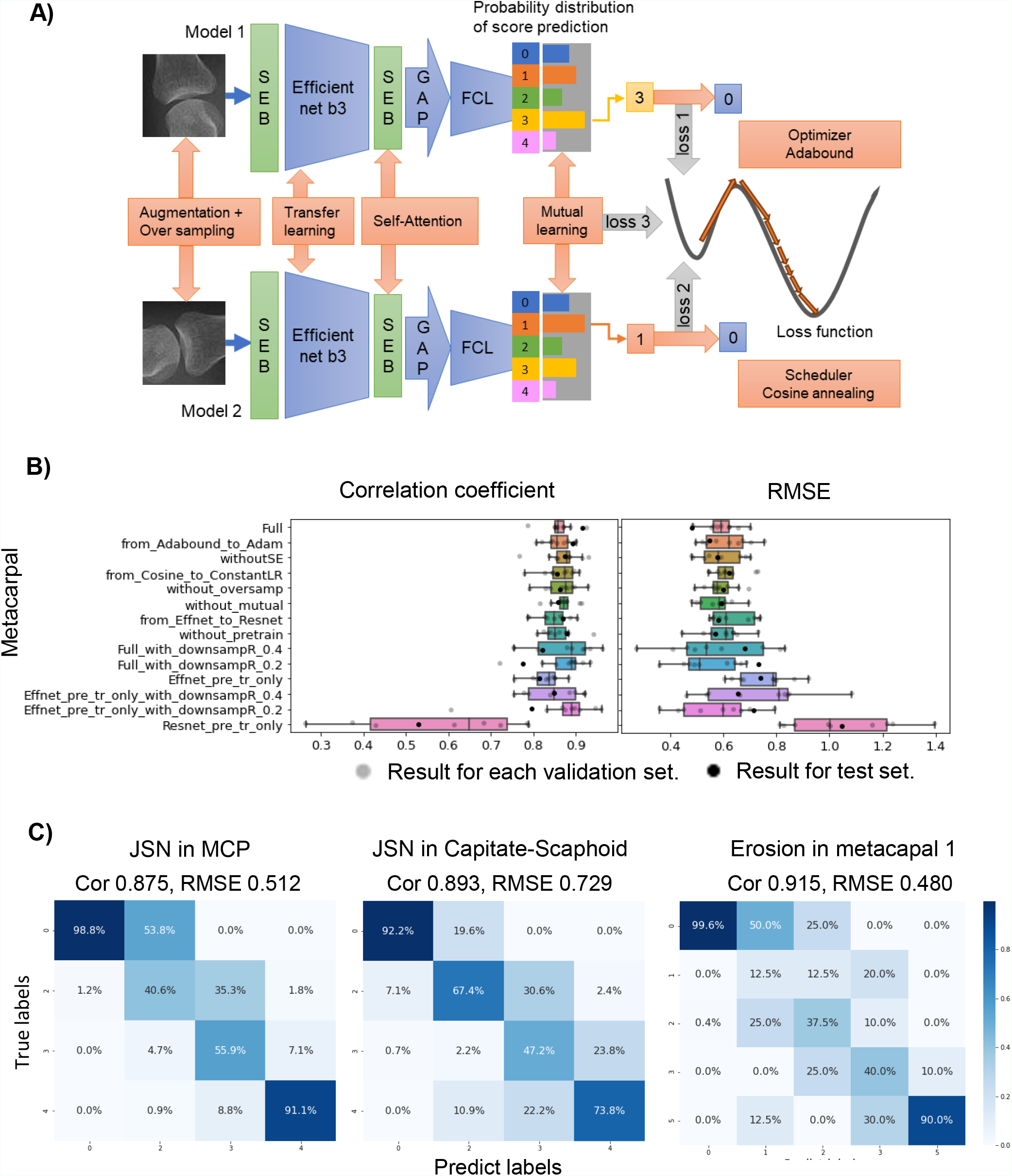
Evaluation of the performance improvement by combining modules. (A) Structure of full model combining all modules. (B) Visualization of combined effect using box plots. Correlation coefficient and Root Mean Squared Error (RMSE) of each model are shown. To compare all models on the same test images, we first divided each joint image dataset into 10 segments, used one as a test set (black dots), and cross-validated the rest (gray dots). (C) Confusion matrix with prediction and true label. ‘Score 4’ in erosion and ‘score 1’ in JSN could not be displayed because few teacher labels were available owing to the constraints of SHS reading. These scores were difficult to train, the model could not regress, and few were included in the test set. See Extended Figure S5 for the score distribution of each joint and constraints of SHS reading. Extended Figure S6 and S7 provide the results for the other joints. Cor, correlation coefficient; FCL, fully connected layer; GAP, global average pooling; JSN, joint space narrowing; MCP, metacarpophalangeal joint; SEB, squeeze-and-excitation block.

**Table 1.** Comparison between correlation coefficient of present and previous models.

|  | Present model | Previous model [2] |
| --- | --- | --- |
| JSN |  |  |
| 2-5PIP | 0.89 | 0.81 |
| 1-5MCP | 0.88 | 0.91 |
| 3-5CMC | 0.86 | 0.84 |
| Around scaphoid bone | 0.87 | 0.88 |
| Erosion |  |  |
| Finger | 0.86 | 0.66 |
| MC1 & Trapezium & Scaphoid | 0.90 | 0.76 |
| Lunate & Ulna & Radius | 0.86 | 0.79 |

The present and previous models constructed in RA2 Dream Challenge [2] differ in the number of joint scores output from each wrist joint image. We calculated the average of the correlation coefficients for the three joints in “MC1&Tra&Sca” and “Luna&Ulna&Radius” and compared them with those in the previous models because the correlation coefficients for each joint have not been reported in the previous models. CMC, carpometacarpal; JSN, join space narrowing; MCP, metacarpophalangeal; MC, metacarpal; PIP, proximal interphalangeal.

We then assessed the combined effects of the modules. We first compared the full model with models with removed or replaced all modules from the full model. Installing all modules in a pretrained EfficientNet model (“Effnet-pre-tr-only” in Figure 3B and Extended Figure S6) and pretrained ResNet model (“Resnet-pre-tr-only” in Figure 3B and Extended Figure S6) substantially improved both the correlation coefficient and root mean square error (RMSE). We then compared the full model with those that removed or replaced one module from the full model. For most joints, the correlation coefficient and RMSE of the full model exceeded those of models that removed or replaced one module (from “from_Adam_to_Adabound” to “without_pretrain” in Figure 3B and Extended Figure S6). Finally, we compared the full model with models trained using the downsampling dataset (“Full_with_downsampR_0.4” and “Full_with_downsampR_0.2” in Figure 3B and Extended Figure S6). Both performance indicators in the validation set tended to worsen in the test set, possibly because of overfitting to the downsampling dataset.

Subsequently, we visualized the regions of the model’s attention using Score-CAM.[19] The model focused on relevant regions for scoring in the joints with lower scores and in higher scores, such as with dislocations and ankylosing (Figure 4). Furthermore, we examined joint images with large differences between predicted and true labels. As shown in the confusion matrix, the percentage of images with a difference of four in JSN and five in erosion was low (Figure 3C, Extended Figure S7). However, our model predicted false positives (FPs) with large differences in estimating the JSN scores for joints similar to pseudo-space narrowing because of hand rotation, although it correctly predicted scores for pseudo-space narrowing joints (Extended Figure S8). The FPs in erosion scores were predicted when rotated hands, rings, or some dislocated/ankylosed joints were present (Extended Figure S8). Moreover, the model predicted false negatives (FNs) with large differences in the JSN scores when joints were misidentified owing to severe deformity and FNs with large differences in erosion scores were predicted when partial ankylosis and collapse existed (Extended Figure S9).

**Figure 4.**
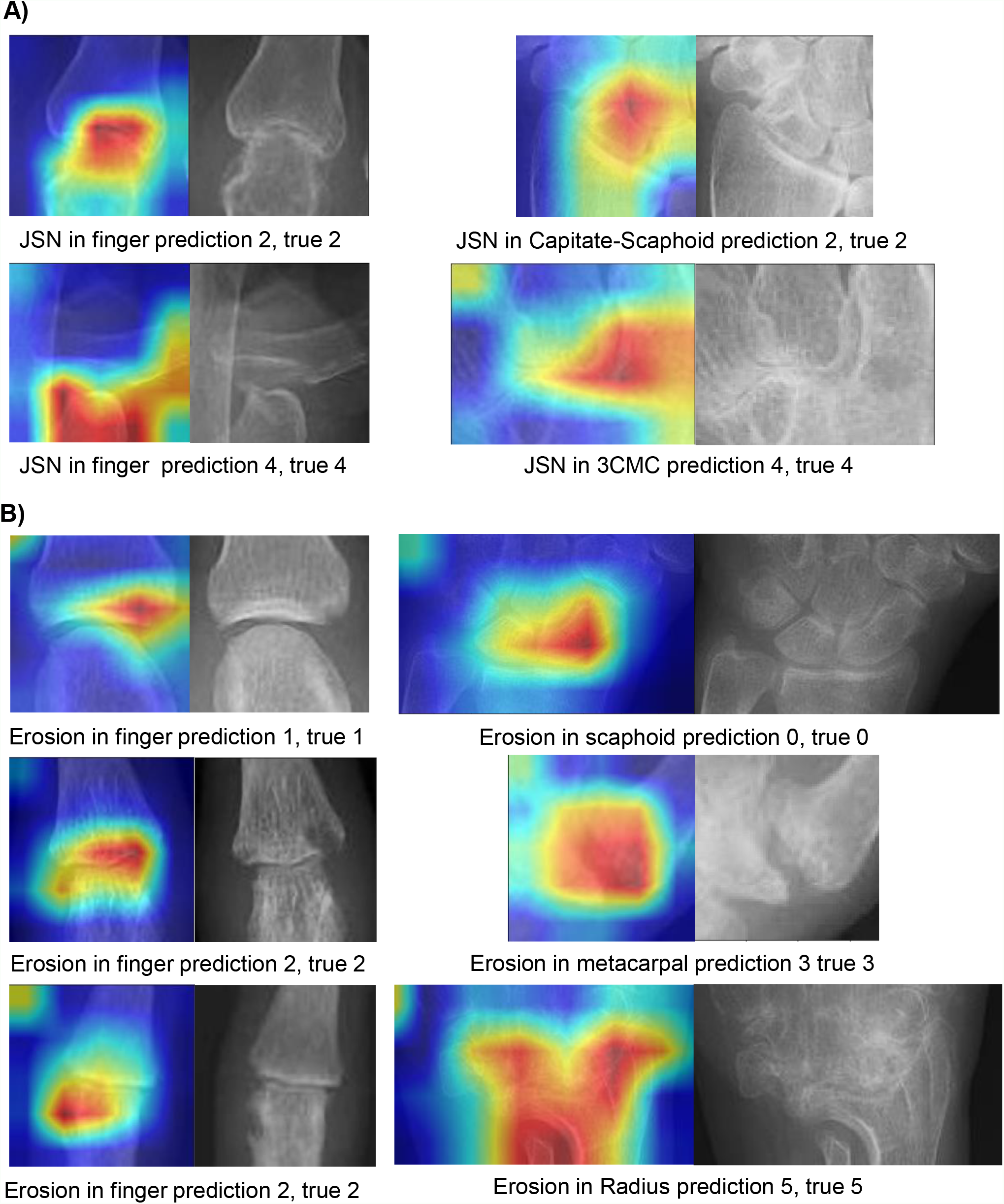
Visualization of the model’s attention. We used score-CAM to visualize the regions of the attention of the model in JSN (A) and in erosion (B). CMC: carpometacarpal joint

Radiographs of the hand in the rotated position may be mislabeled because the joint space appears narrower (Extended Figure S8). Therefore, to examine the effects of hand rotation on the scores, we extracted 770 pairs of frontal and oblique hand radiographs taken on the same day and estimated the angle formed by the straight line connecting the 2MCP and 5MCP. Finally, we regressed the angle and score difference per joint between the frontal and oblique images (Supplementary Methods). From the results of the regression, the variation in score per finger joint was less than 0.5 and that per wrist joint was less than 1.0 when the rotation angle was less than 20° (Extended Figure S10).

### Verification of applicability to clinical research

Thus far, we have constructed a high-performance model. However, we must confirm that this model can replicate previously reported findings derived from the scores of expert readings. Therefore, we selected 1,503 patients with hand radiographs taken from at least two different time points from INDRA and stratified the progression of joint destruction according to anti-cyclic citrullinated peptide antibodies (anti-CCP) and the rheumatoid factor (RF). As in previous reports,[20] the ΔSHS scored by our full model was well differentiated based on stratification by positivity of the serological markers (Figure 5A); however, with Effnet-pre-tr-only-model, the differences in scores between groups decreased; that is, the resolution degraded (Figure 5B).

**Figure 5.**
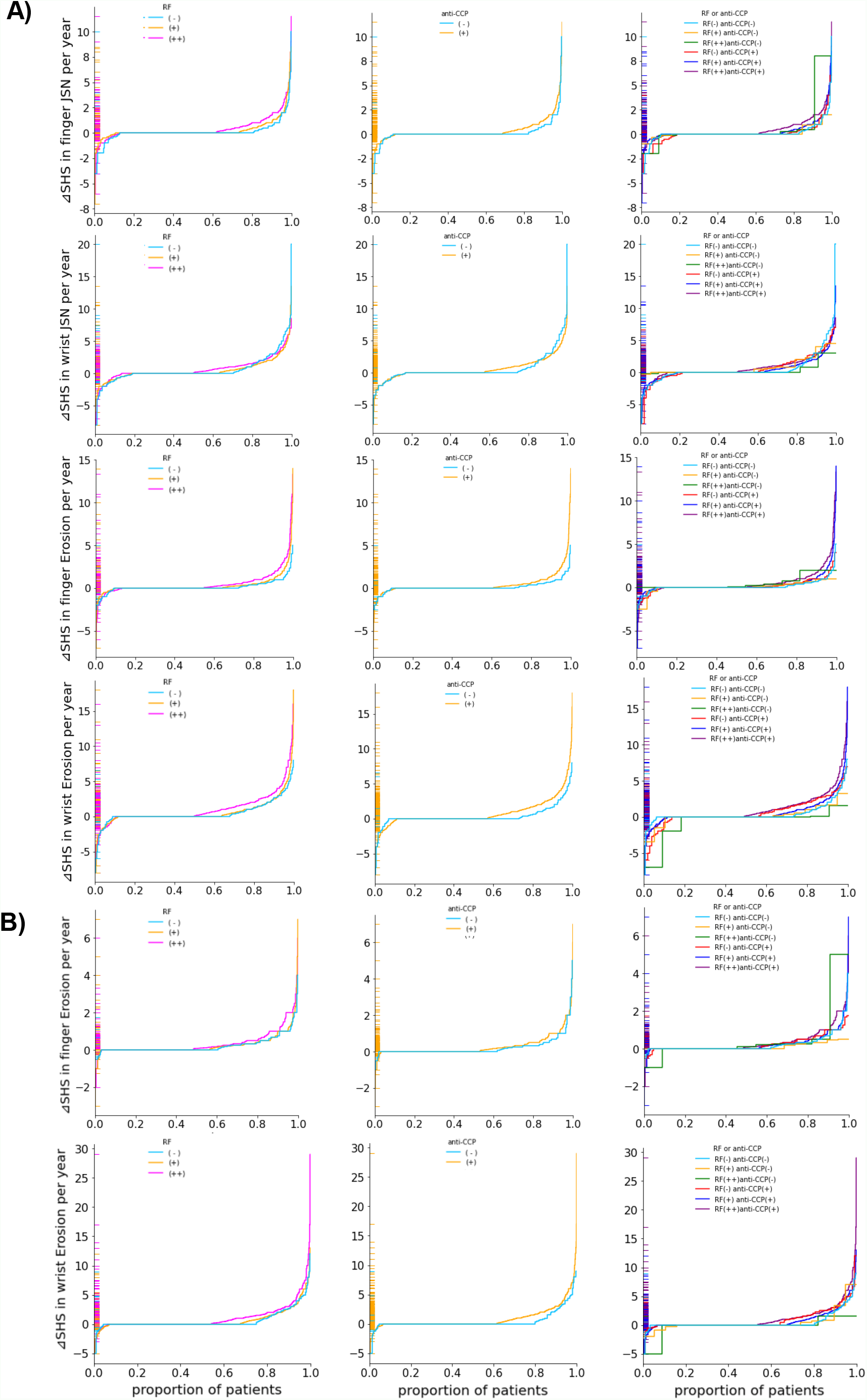
Subgroup analysis for replicating the association between the progression of joint destruction and autoantibody positivity. We performed a subgroup analysis of the JSN and erosion scores stratified by RF values and anti-CCP positivity, and a combination of both. We used the scores predicted by the full model (Panel A) and pretrained-only Efficientnet model (Panel B). We extracted 1503 patients with radiographs taken from at least two time points and calculated the ΔSHS per year, divided by the interval between the radiographs. For stratification by RF, we divided the patients into three groups according to the values as follows: negative/-; ≤ 15, weak positive/+; 15 >, ≤ 100, and strong positive/++; > 100. CCP, cyclic citrullinated peptide; RF, rheumatoid factor; SHS, Sharp/ van der Heijde scores.

Next, to characterize the SHS distribution, we scored the hand radiographs of 4,264 patients within INDRA using our model and performed hierarchical clustering. Consequently, patients in INDRA were divided into eight clusters (Figures 6A and 6B). To test whether these clusters were clinically important, we performed a subgroup analysis stratified by cluster. We demonstrated that the wrist erosion score was more likely to progress in patients classified in clusters 4, 7, and 8, whereas the finger erosion score was more likely to progress in patients classified in clusters 1 and 3 (Figure 6C and Extended Figure S11). Therefore, we extracted patients classified in clusters 1, 3, 4, 7, and 8 and further stratified them by their anti-CCP and RF statuses and determined that they could be further differentiated (Extended Figure S12). Finally, we performed a multivariable analysis with multiple regressions to confirm whether these clusters were independent of clinical factors. In addition to known factors, clusters 4, 7, and 8 were independent risk factors for wrist and finger erosion progression, whereas clusters 1 and 3 were independent risk factors for finger erosion only (Supplementary Tables 1 and 2).

**Figure 6.**
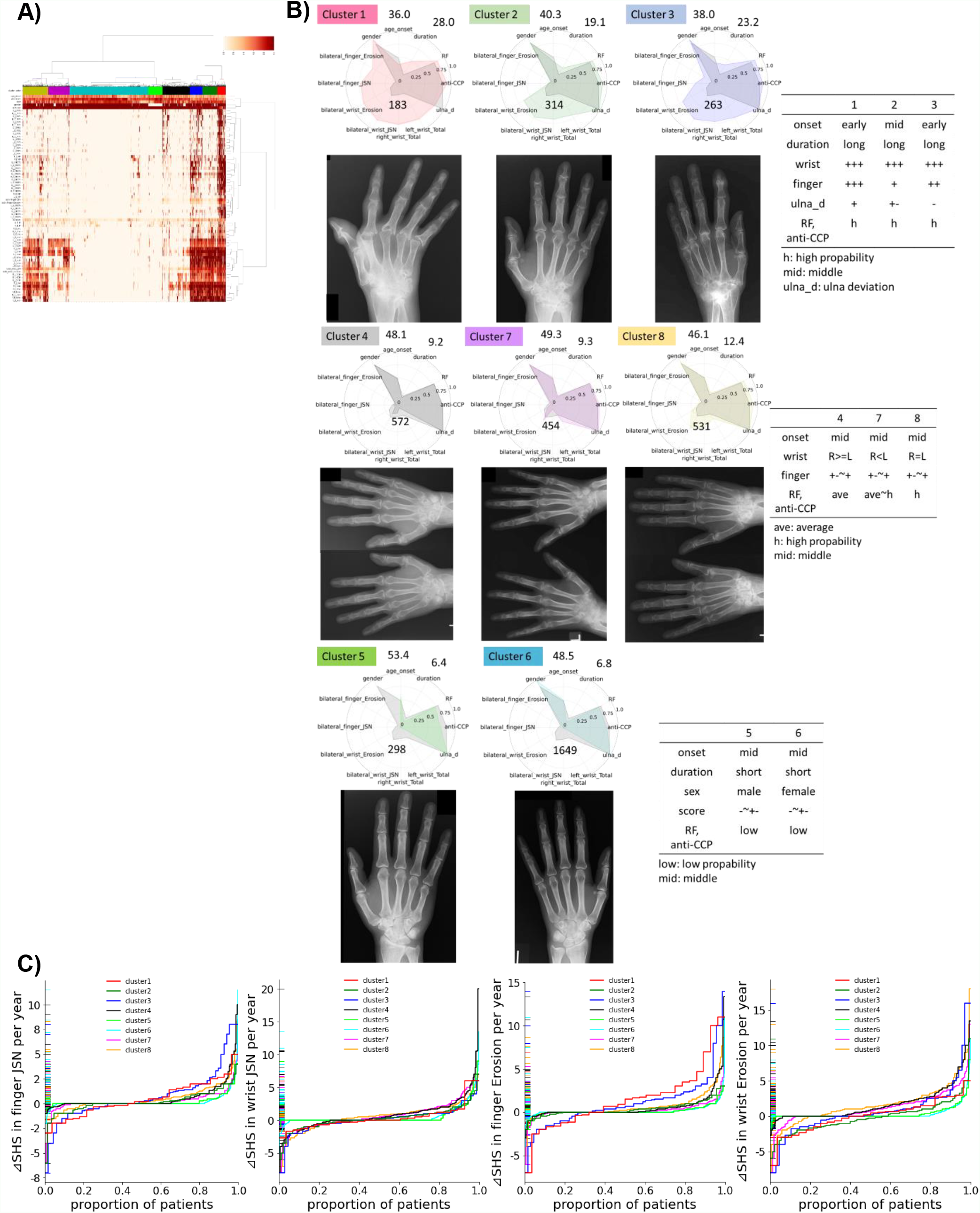
Overview of the results of hierarchical clustering. To characterize the largest SHS distribution, we performed hierarchical clustering (A) and depicted radar charts to clarify the difference between each cluster after adding information on serological markers (B). The gray areas in the radar charts indicate the average of all patients. The mean values of the normalized variables are shown in the radar chart between 0 and 1. The onset classification was defined as follows: early onset, ≤ 40 years; middle-onset, > 40 years; ≤ 60 years, late-onset; and > 60 years. We then conducted a subgroup analysis of the radiographic progression of JSN and erosion in the finger and wrist according to clusters (C). ave, average; CCP, cyclic citrullinated peptide; h, high; JSN, joint space narrowing; low, low probability; mid, middle; RF, rheumatoid factor; SHS, Sharp/ van der Heijde scores; ulna_d, ulnar deviation.

### Construction of the prediction model for radiographic progression

Finally, we constructed a model to predict radiographic progression using a GCN. In brief, we connected patients to form a graph structure by cosine similarity based on the feature vectors (explanatory variables) and classified unlabeled patients in the network based on both the graph structure and feature vector using the GCN (Figure 7A, Supplementary Methods). We performed 10-fold cross-validation and compared the average area under the curve (AUC) of the GCN on predicting joint destruction to that of RF-positive, anti-CCP-positive, and random forest with and without Boruta (Supplementary Methods)[21]. The AUCs for predicting finger and wrist erosion in the GCN model were 0.800 and 0.748, respectively, which are superior to those for the other models (Figure 7B and Supplementary Table 3).

**Figure 7.**
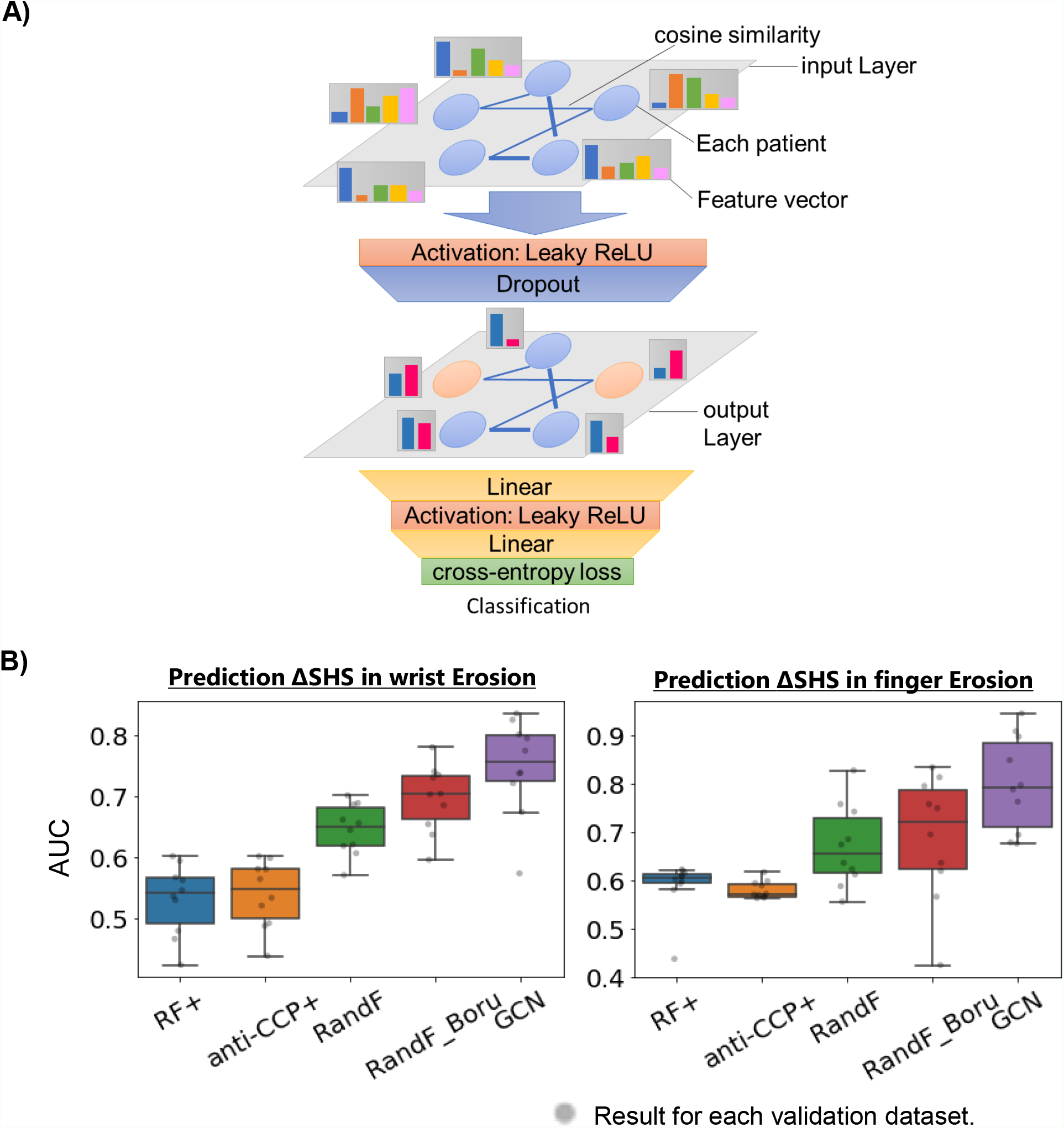
Construction and results of the prediction model for radiographic progression. We constructed a prediction model using GCN (A) and compared the model with other serological markers and random forest with and without Boruta through 10-fold cross-validation (B). The AUC for predicting the progression of finger and wrist erosion is depicted using box plots (B). Boru, Boruta; CCP, cyclic citrullinated peptide; GCN, graph convolutional network; RandF, random forest; RF, Rheumatoid factor.

We calculated the feature importance (Supplementary Methods) in the random forest and GCN models and extracted the top 10 features. The binarized clusters were the top features in predicting wrist erosion but not in predicting finger erosion. (Supplementary Tables 4 and 5).

## DISCUSSION

This study is the first to demonstrate that a CNN-based automated scoring model for joint destruction can be applied to clinical research. We demonstrated that the model with the combined module had better performance than the model without the combined module and that this improvement in performance is clinically meaningful by evaluating the results of the subgroup analysis with the RF and anti-CCP for radiographic progression of joint destruction. Furthermore, we are the first to report that the GCN model is superior to previous machine learning methods in predicting joint destruction.

Our success in creating an automated scoring AI system is derived from three main factors. First, we collected many hand radiographs with high SHSs before the current standard of treatment was initiated. In Japan, methotrexate was approved in 1999 and infliximab (the first biologic agent approved in Japan) in 2003. We collected SHS data and with radiographs since these years. Although our data were imbalanced (Extended Figure S5), we constructed a model with a dataset that included a more patients (number of patient train: 1074, validation: 134, test: 134) and radiographs with highly progressed joint destruction than the previously reported dataset with 367 patients in the training set.[2] Second, weakly supervised learning using U-Net can accurately detect the coordinates of radiographs with severe joint destruction. The detection of wrist joints with progressive destruction is a particularly challenging task,[3] and most reports on AI scoring models are limited to finger score predictions.[4,22,23] ResNet34, used in a previous report,[2] could not achieve highly accurate detection in our hand images, possibly because our dataset contains more severe joint destruction. In addition, a CNN that directly regresses landmarks, such as joint coordinates, is considered a model that performs a highly nonlinear transformation from images to coordinates; thus, accurately estimating coordinates is difficult.[24] Weakly supervised learning with heatmap regression allows image-to-image mapping to reduce the complexity of the nonlinear transformations. The U-Net for heatmap regression was sufficiently accurate for identifying the joint coordinates. Third, our team was composed of clinicians who incorporated their perspective into the model. For example, a previously reported model, which was constructed by a team without clinicians, predicted scores as a regression problem (i.e., the output was a continuous value). This method improves accuracy when training on small datasets, but from a clinician’s perspective, JSN scores of three and four cannot be expressed as continuous values because they are defined by the presence or absence of JSN *or* dislocation. We believe that a team that includes clinicians can construct an optimal module.

Our new findings showed that GCN improves the prediction of joint destruction and that clusters are an important predictor of bone erosion. GCNs have achieved remarkable results in compound estimation and analysis of omics data.[25,26] However, no clinical applications in rheumatology have been reported to date. The reason for the high accuracy of the GCN is believed to be that it uses features (age, gender, etc. in our model) as in conventional models, as well as their graph structure for estimation.[26] The clustering results were an independent risk factors and an important feature for predicting bone erosion (Table 4), which may replicate the previously reported evidence of the presence of onset joint destruction predicting future bone erosion.[27,28] However, as similar findings were obtained using AI without arbitrary, manual selection is considered a novel and important finding.

A limitation of this study is that we could not perform external validation owing to the lack of an available image dataset with SHSs. Therefore, training, validation, and testing were performed using image data from the same cohort. The prediction accuracy may be degraded if a discrepancy exists between our test set and the data in a clinical setting. However, our full model could replicate the known findings of the association between radiographic progression and serological markers, which were excluded in the AI construction process. Furthermore, in a subgroup analysis using the SHSs predicted by the full and base CNN models, the resolution of the full model was better than that of the base CNN model. Therefore, we believe this evidence will at least mitigate the criticism of the models being inapplicable in real-world practice because of the overfitting to a particular dataset. Another limitation is that our model does not score by comparing with previous images, as in expert scoring. Although the hand rotation examined in this study and joint deformities may cause overestimation, relative evaluation by comparison with previous images in expert scoring can minimize it. Therefore, to evaluate the inter-rater reliability of ΔSHS between the relative evaluation by experts and absolute evaluation by our model, we prepared 50 new hand radiographs for 25 patients taken twice (total hand SHSs from a reading expert, median 24 [minimum 0, interquartile ranges 2-69, maximum 242]) and calculated the ICC to be 0.75 (95%, confidence interval of 0.51–0.88). This value was inferior to previously reported inter-professional ICCs (0.83–0.86),[29,30] but remained high. Most importantly, our model replicated known findings and has applicability to research, although improving the accuracy remains a challenge.

In conclusion, we constructed a high-performance model combined with various modules for joint score prediction, which replicated previously reported findings and characterized the largest SHS distribution calculated using this model. Furthermore, we demonstrated, for the first time, that the GCN is useful in the field of clinical rheumatology. We believe that this model is a promising tool for future studies, such as in GWAS for joint destruction, which includes thousands to millions of cases that are difficult to realize with human scoring.

## Data Availability

The data using analysis that support the findings of this study are available on reasonable request to the authors.
The imaging data are not publicly available due to restrictions (containing information that could compromise the privacy of research participants)
Code is available at https://github.com/honda-s691470/SHS_NET/ 　and https://github.com/honda-s691470/GCN_SHS_NET

## Contributors

K.I. conceived the study design. S.H. conducted all analyses with the help of K.I., and M.H. and S.H. wrote the manuscript. K.I., S.H., E.T., and M.H. collected samples and clinical information. All authors reviewed and approved the final manuscript.

## Acknowledgment

We thank all patients in the IORRA database and all members of the Institute of Rheumatology, Tokyo Women’s Medical University Hospital, for the successful management of the IORRA study cohort.

## Ethics approval

The IORRA cohort studies (#2952-R and #2922-R16) were approved by the ethics committee of Tokyo Women’s Medical University, and informed consent was obtained from all patients before each survey.

## Patient and public involvement

Patients and/or the public were not involved in the design, conduct, reporting, or dissemination of this research.

## Patient consent for publication

Not required.

## Data sharing statement

The data using analysis that support the findings of this study are available on reasonable request to the authors.

The imaging data are not publicly available due to restrictions (containing information that could compromise the privacy of research participants)

## Code availability

Code is available at https://github.com/honda-s691470/SHS_NET/ and https://github.com/honda-s691470/GCN_SHS_NET

