## Extended for "Development of a scoring model for the Sharp/van der Heijde score using convolutional neural networks and its clinical application in predicting radiographic progression using a graph convolutional network"

### Extended figures

**A)**

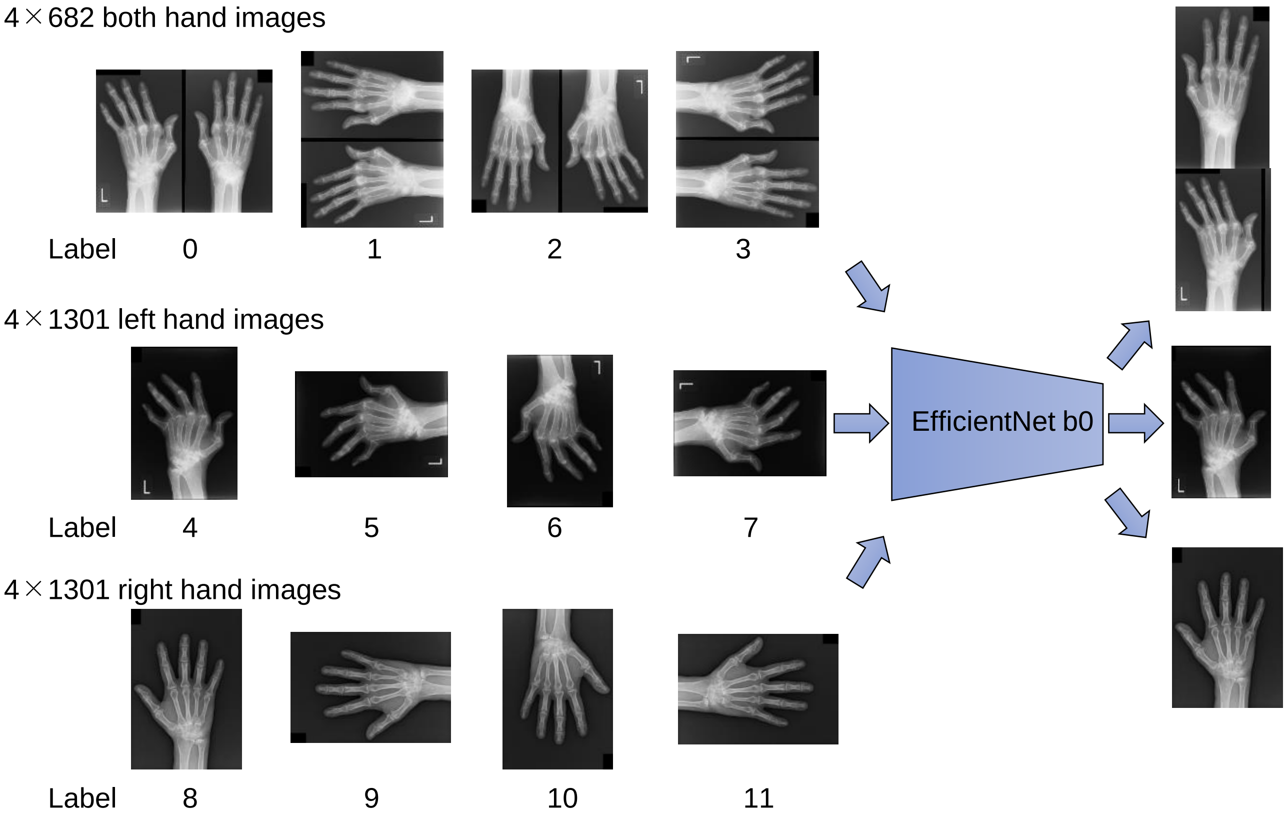


**B)**

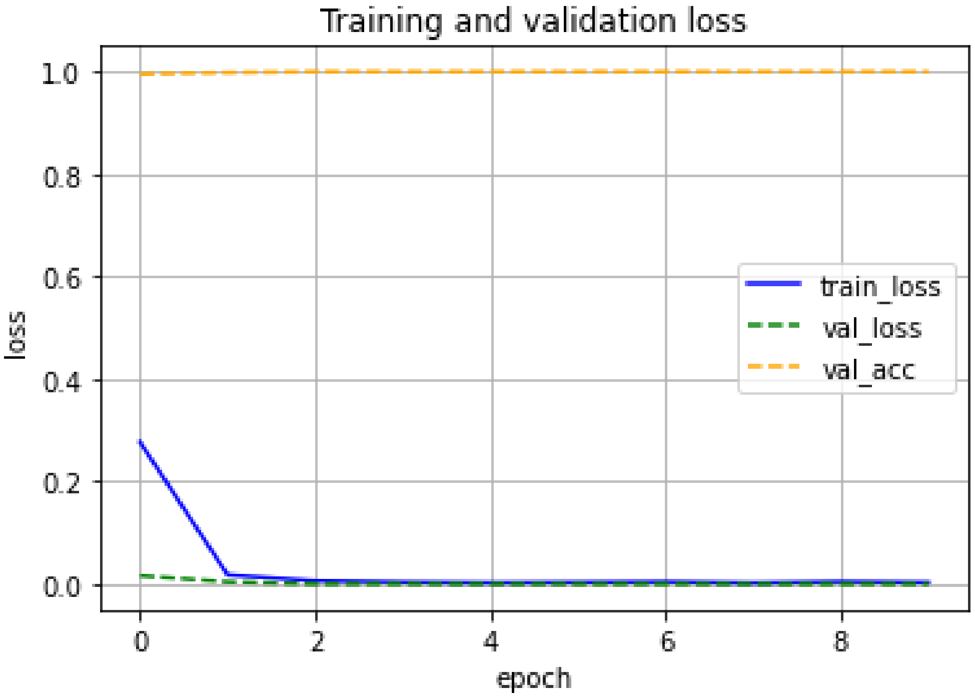


**Figure S1. Overview and result in the orientation phase.**
In the orientation phase, a total of 12,866 images were used. Depending on the image type, labels were created using one-hot encoding as shown in the figure. These images were divided in a ratio of 3:1 (9,852 vs 3,284 images), and the pre-trained EfficientNet b0 was trained and evaluated (A). We depicted Learning curves with train loss, validation loss and validation accuracy per epoch (B). The accuracy of our model reached 100% after 3 epochs.

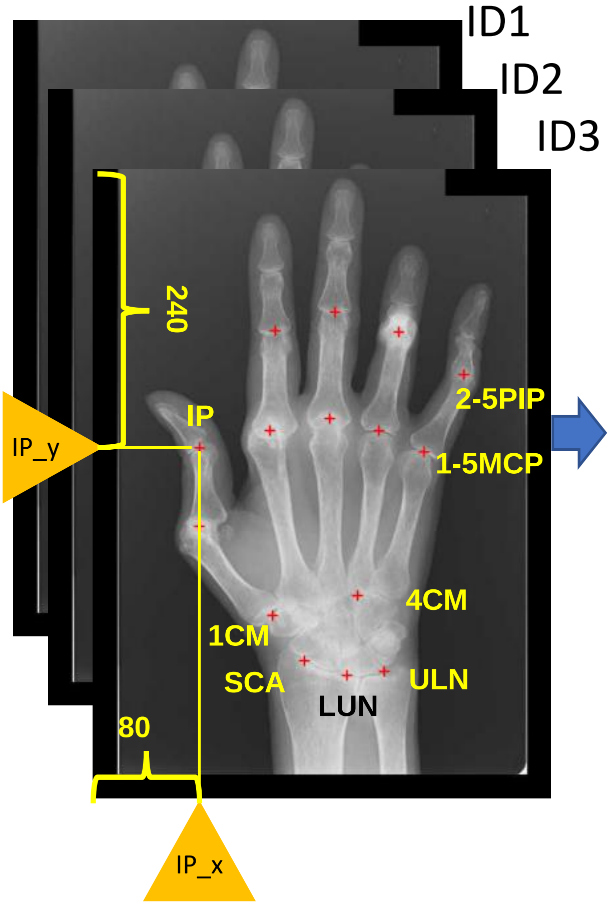


| ID | IP_x | IP_y | PIP2_x | … | SCA_y |
|---|---|---|---|---|---|
| 1 | 80 | 240 | 150 | … | 482 |
| 2 | 116 | 375 | 211 | … | 663 |
| 3 | 538 | 433 | 433 | … | 771 |
| 4 | 72 | 365 | 165 | … | 713 |
| … | | | | | |
| 1381 | 453 | 216 | 542 | … | 847 |

**Figure S2. Obtaining the coordinates of the landmarks in the hand radiographs.**
We first created an in-house script to get the coordinates of the clicked location in the hand radiographs. We then retrieved the coordinates of 15 landmarks from each radiograph as shown in this figure, for a total of 20,715 ground-truth coordinates from 1,381 hand radiographs.
CMC, carpometacarpal joint; IP, interphalangeal joint; LUN, Lunate bone; MCP, metacarpo phalangeal joint; PIP, proximal interphalangeal joint; SCA, Scaphoid bone; ULN, Ulna.

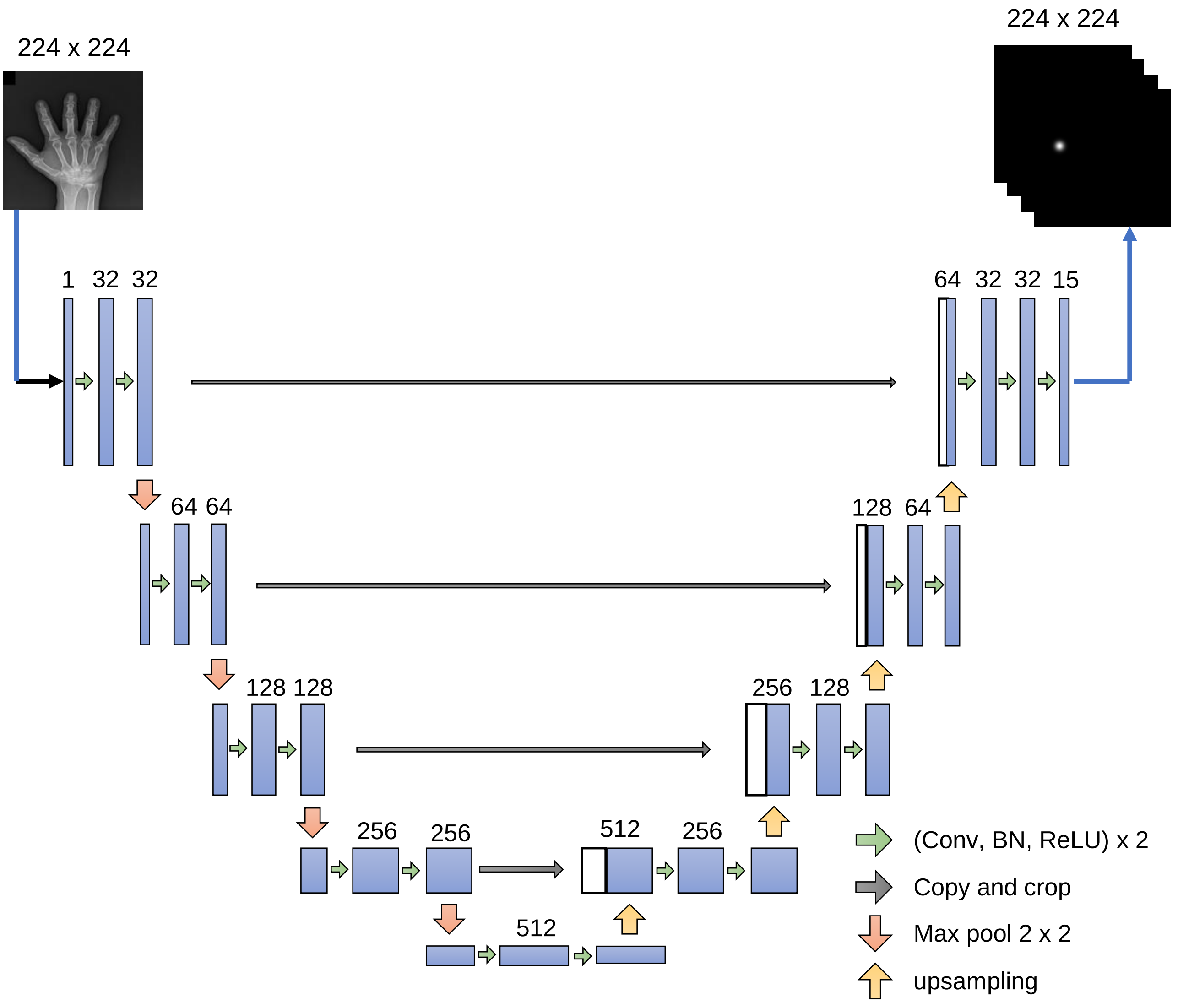


**Figure S3. U-Net structure used in joint detection phase.**
We constructed U-Net base model for joint detection phase. We input the hand radiographs into this model after resizing it to 224x224 pixels. The number of channels is shown above the blue box indicating the multi-channel feature map. The white boxes are feature maps copied from the opposite path and concatenated. The arrows stand for the different operation.
BN, batch normalization.

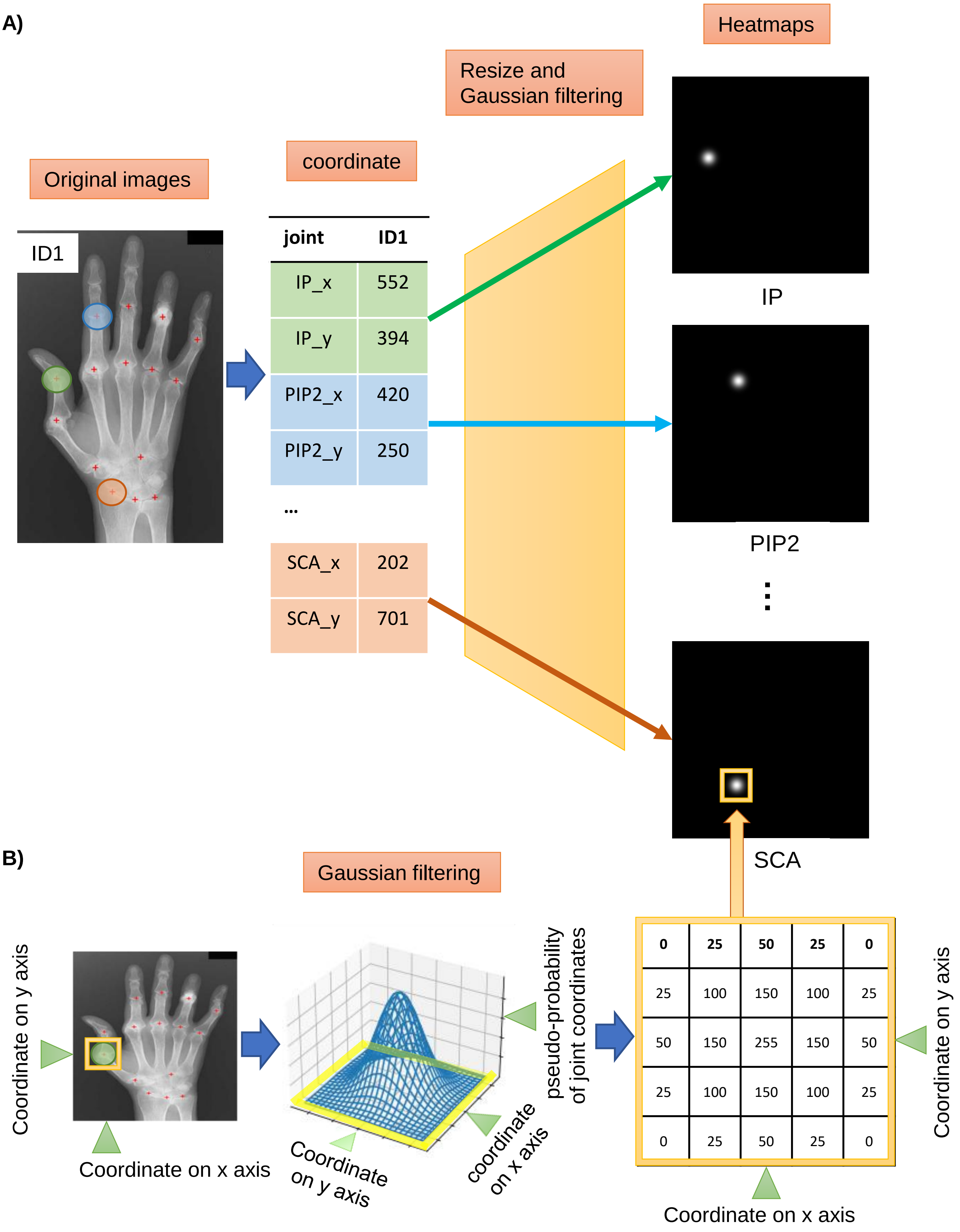


**Figure S4. Preparation for heatmap regression using U-Net.**
We obtained the coordinates of each joint from the original hand graphs using an in-house script, and then generated heatmap images for each joint coordinate using Gaussian filtering (A). Landmark's coordinates (x,y) are passed through a Gaussian filter, which produces an image with increasing values closer to the correct coordinates according to Gauss distribution (B). See supplementary methods for details.
IP, interphalangeal joint; PIP, proximal interphalangeal joint; SCA, scaphoid.

A)

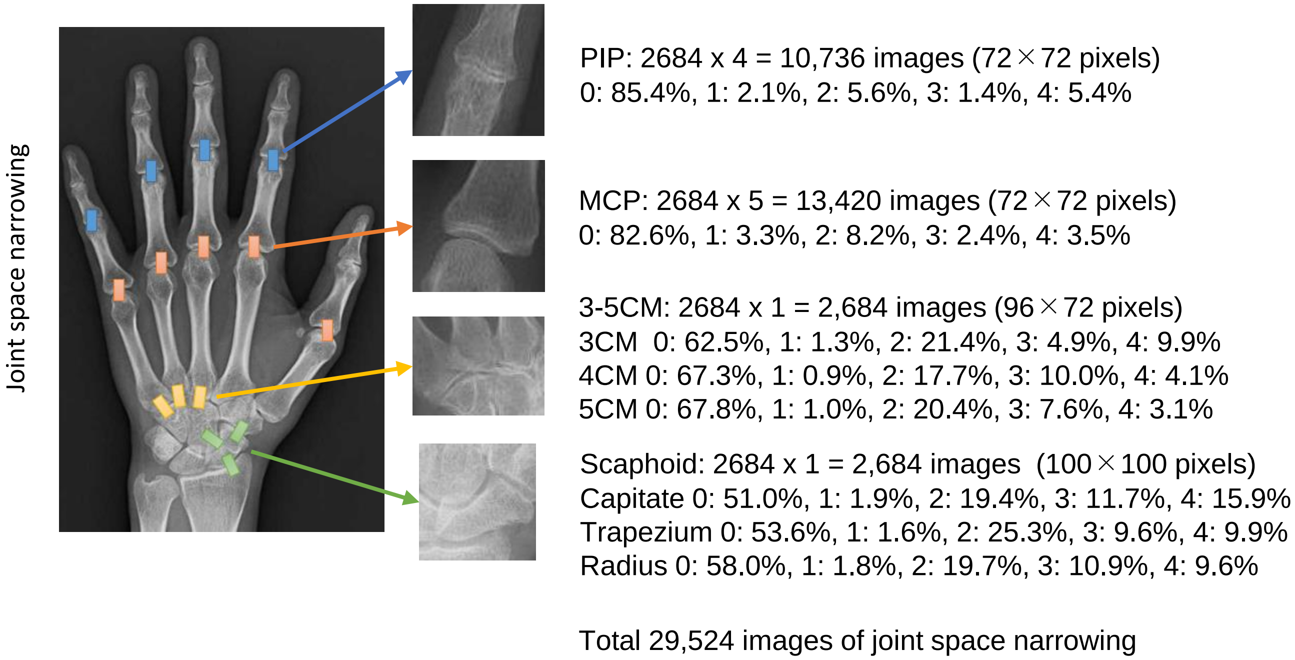


B)

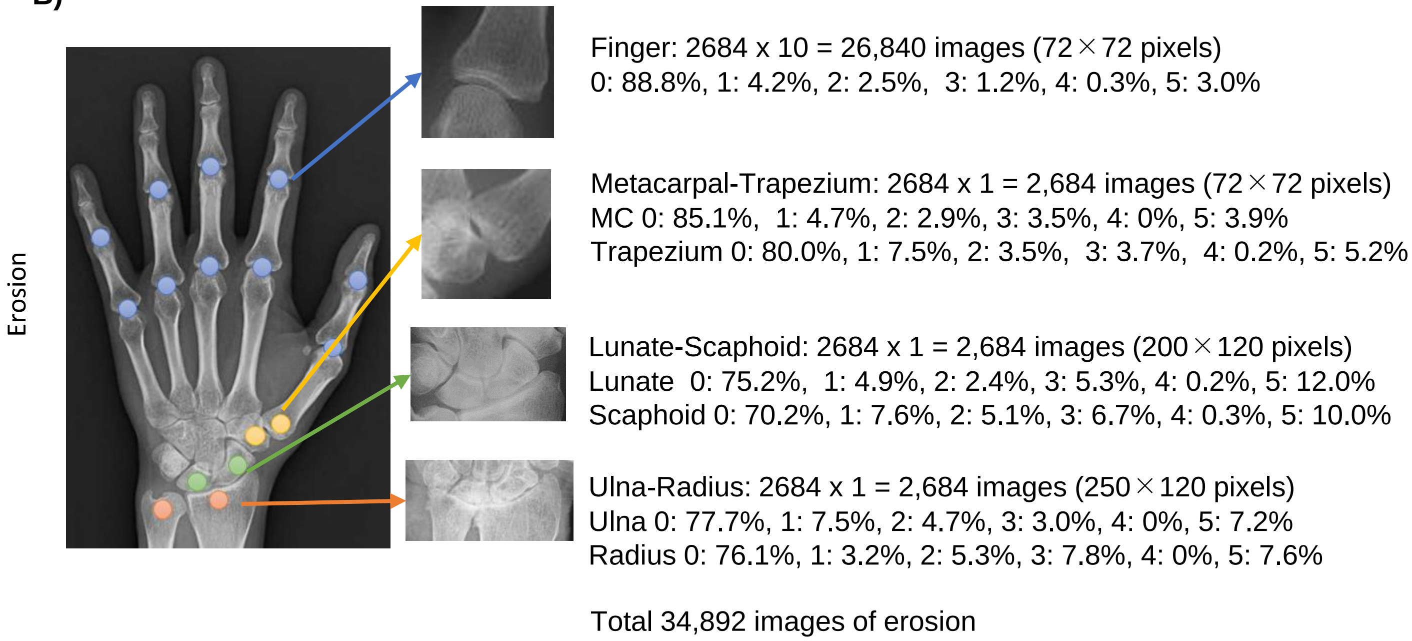


**Figure S5. Number of cropped images and score distributions**
We cropped the images used in the JSN (Panel A) and erosion (Panel B) score prediction using a U-Net based model constructed in a joint detection phase as shown in this figure. The right side of each panel shows the percentage of joint scores. With respect to the JSN score for the SHS method, there is a constraint that a score of 1 should be given as little as possible, and the percentage of the score was low. For an Erosion score of 4, the sum of the Erosion scores of the bones forming the joint must be exactly 4. For example, if bone 1 has a score of 1 and bone 2 has a score of 3, the score is 4. If the score for bone 1 is 3 and the score for bone 2 is also 3, the maximum score is 5 instead of 6 (ceiling effect). Thus, the percentage of joints with a score of 4 is low.

CM, carpometacarpal joint; MCP, metacarpophalangeal joint; PIP, proximal interphalangeal joint.

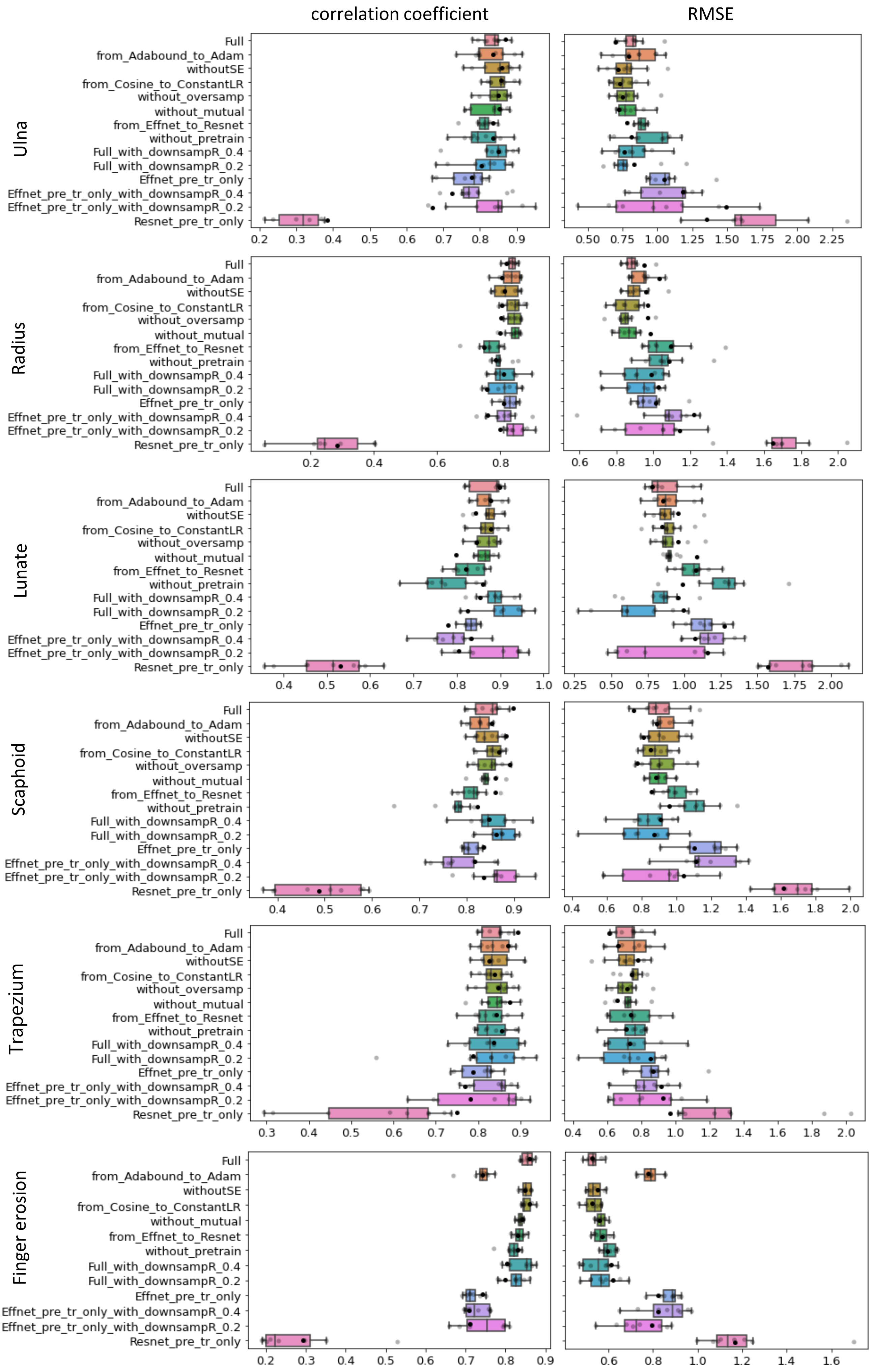

correlation coefficient
RMSE
Ulna
Radius
Lunate
Scaphoid
Trapezium
Finger erosion
Full
from_Adabound_to_Adam
withoutSE
from_Cosine_to_ConstantLR
without_oversamp
without_mutual
from_Effnet_to_Resnet
without_pretrain
Full_with_downsampR_0.4
Full_with_downsampR_0.2
Effnet_pre_tr_only
Effnet_pre_tr_only_with_downsampR_0.4
Effnet_pre_tr_only_with_downsampR_0.2
Resnet_pre_tr_only

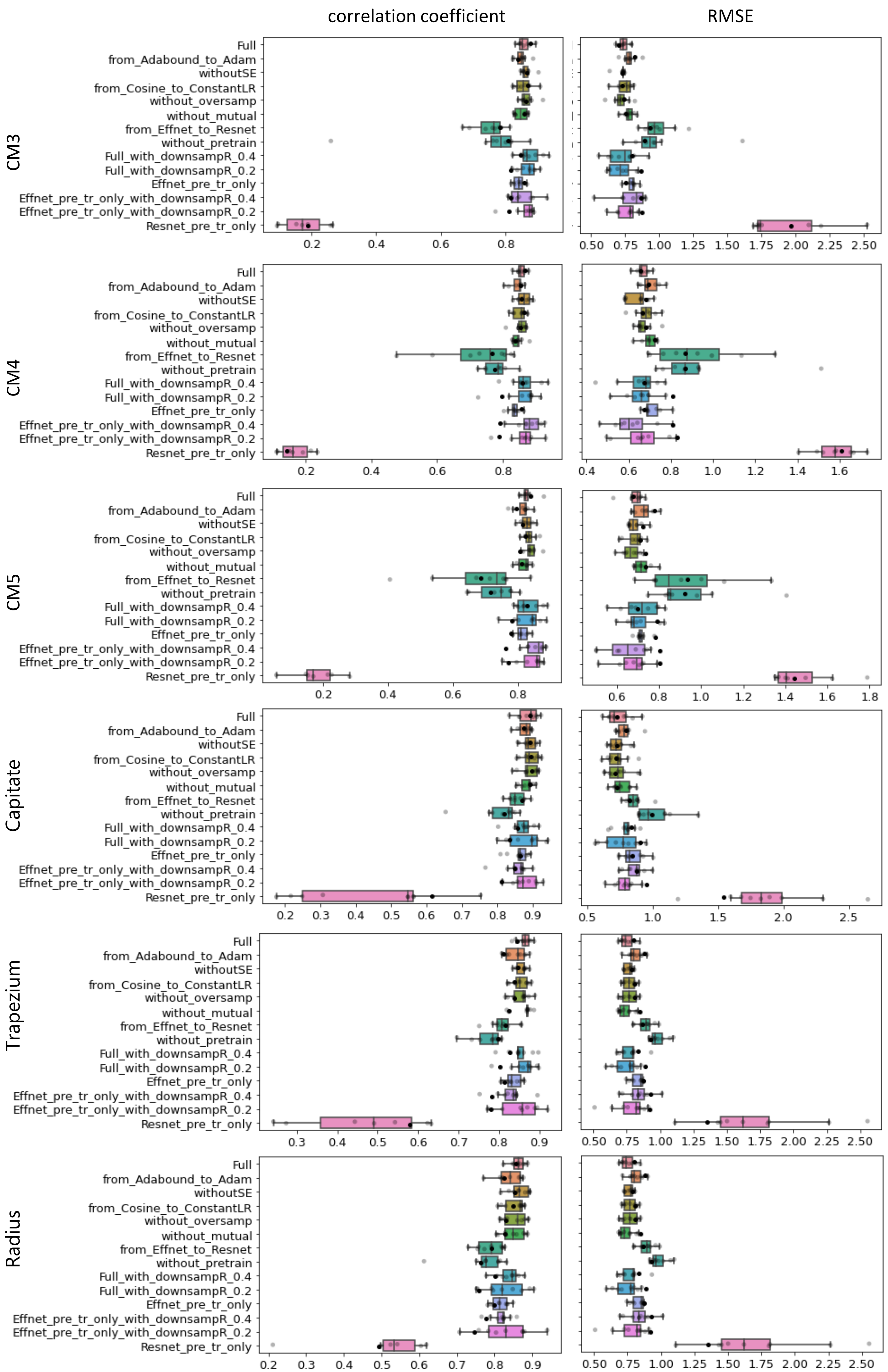

correlation coefficient
RMSE
CM3
CM4
CM5
Capitate
Trapezium
Radius
Full
from_Adabound_to_Adam
withoutSE
from_Cosine_to_ConstantLR
without_oversamp
without_mutual
from_Effnet_to_Resnet
without_pretrain
Full_with_downsampR_0.4
Full_with_downsampR_0.2
Effnet_pre_tr_only
Effnet_pre_tr_only_with_downsampR_0.4
Effnet_pre_tr_only_with_downsampR_0.2
Resnet_pre_tr_only

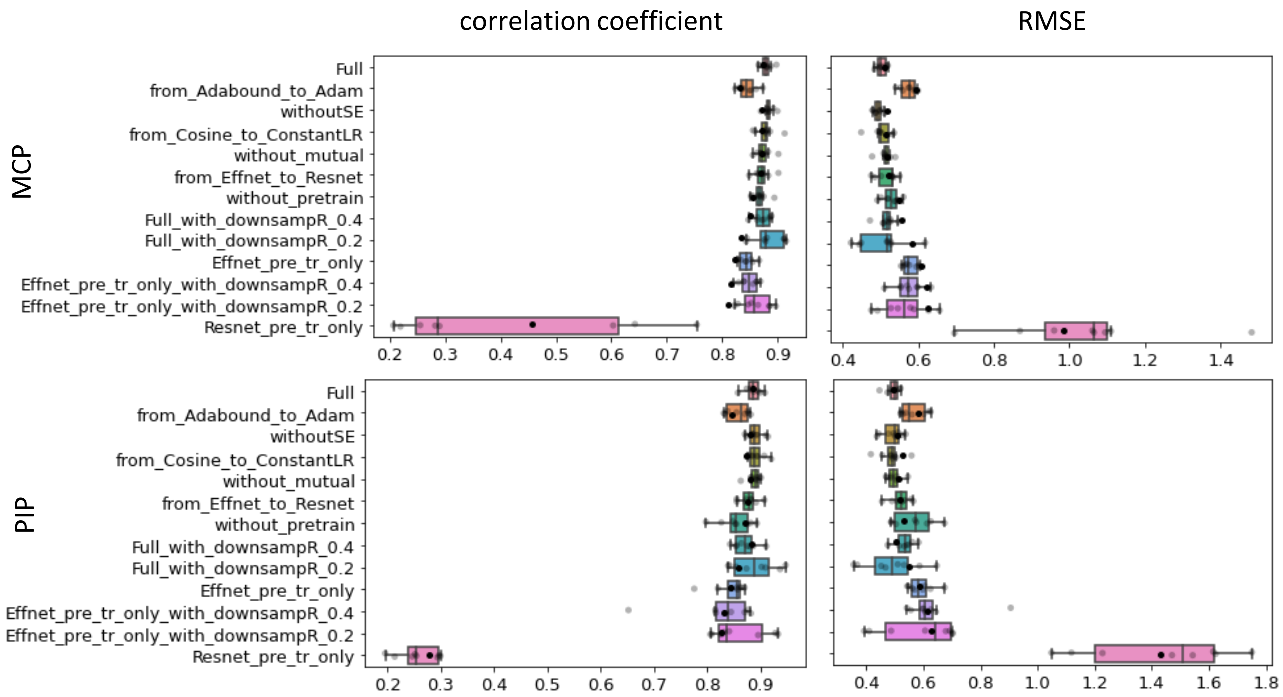


**Figure S6. Performance comparison between Full model and other models at all joints.** Unlike the wrist joints, model with oversampling had lower performance for finger erosion, PIP and MCP in JSN. The number of image data for the wrist joint differs from that for the finger joint by a factor of up to 10, which may be caused by the obvious difference in data structure. For this reason, Oversampling was not used for the finger joints.

CM, carpometacarpal joint; MCP, metacarpophalangeal joint; PIP, proximal interphalangeal joint; RMSE, root mean square error.

A)

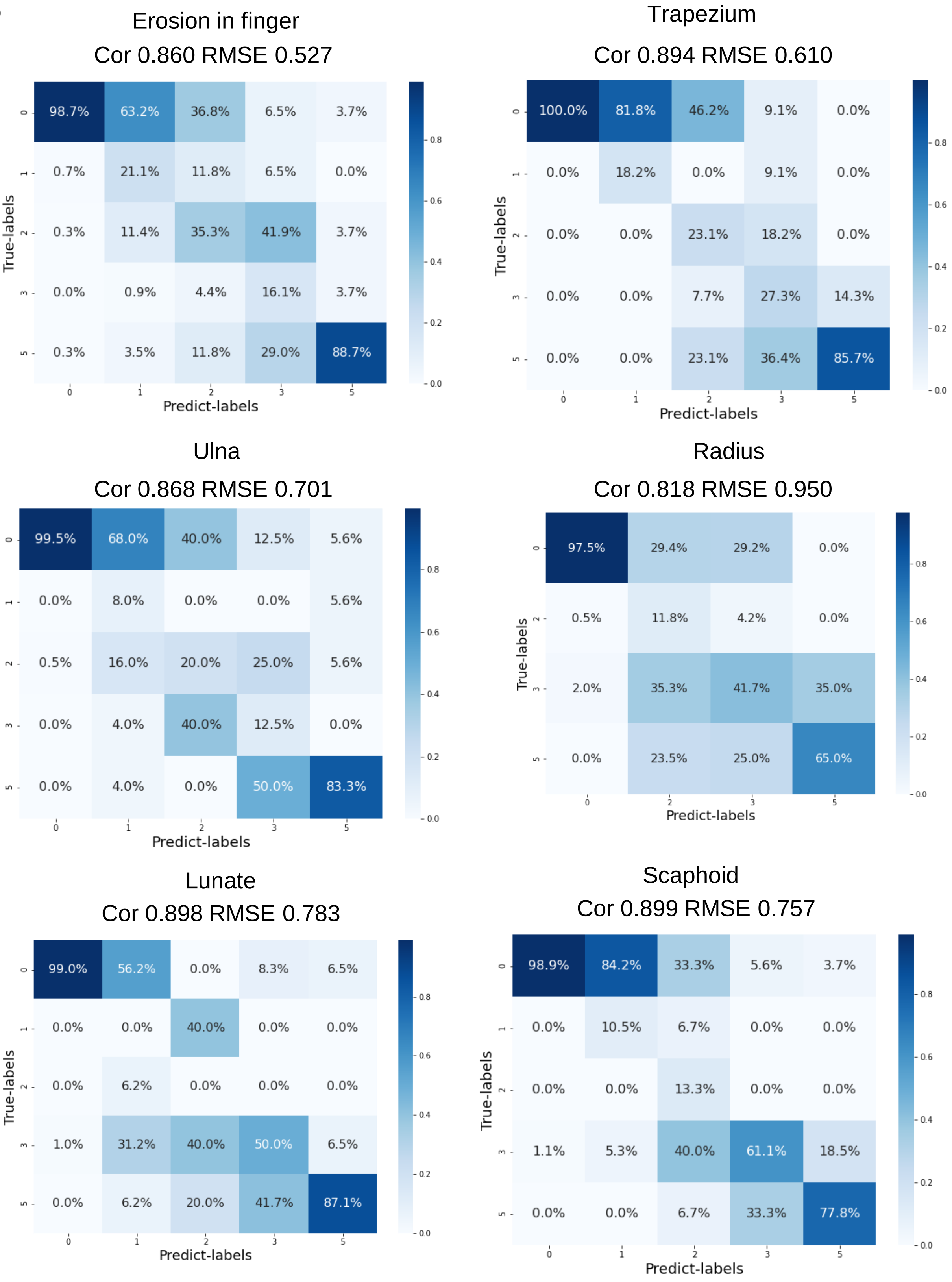

Erosion in finger
Cor 0.860 RMSE 0.527
Trapezium
Cor 0.894 RMSE 0.610
Ulna
Cor 0.868 RMSE 0.701
Radius
Cor 0.818 RMSE 0.950
Lunate
Cor 0.898 RMSE 0.783
Scaphoid
Cor 0.899 RMSE 0.757
True-labels
Predict-labels

B)

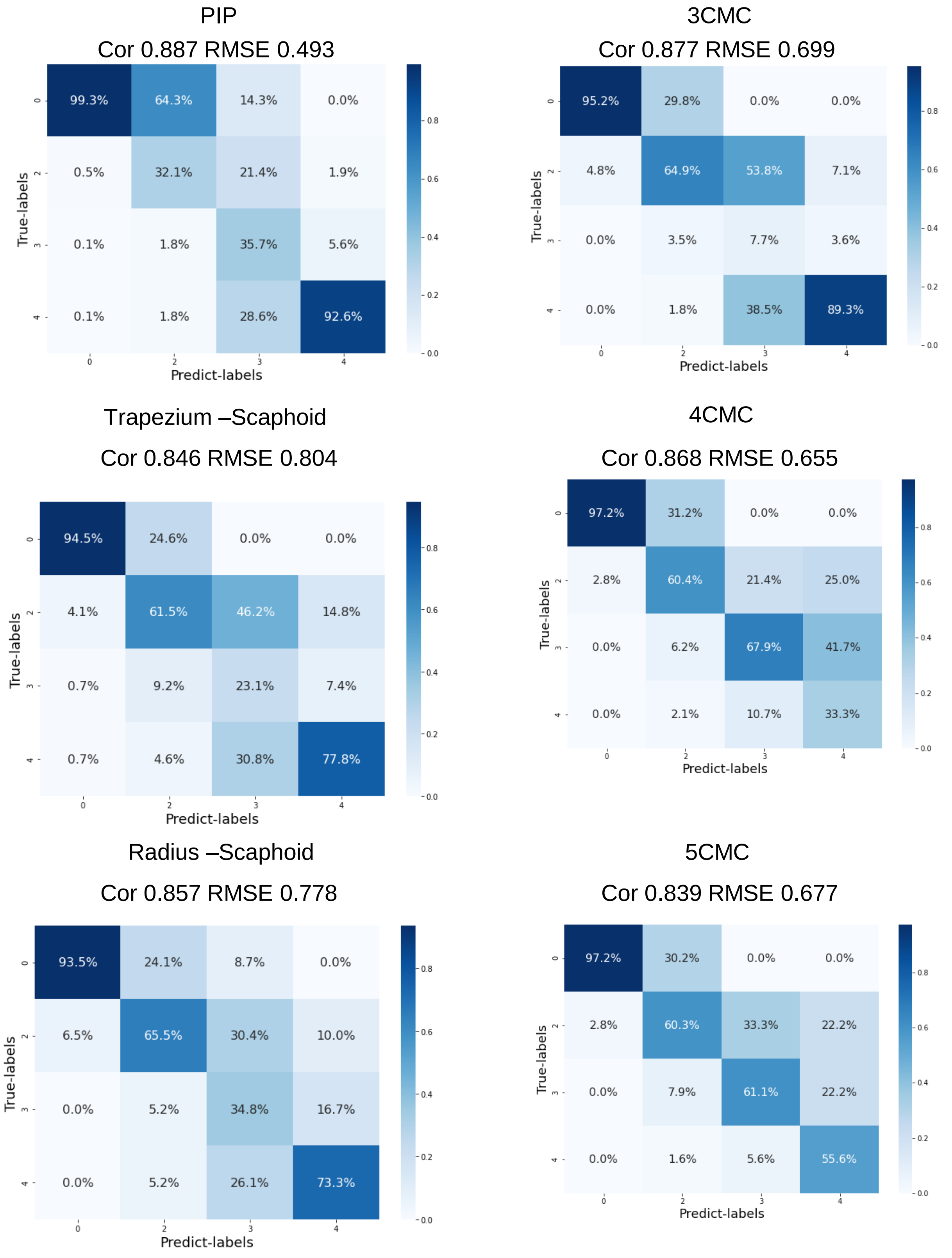


**Figure S7. Confusion matrix with Prediction and True label.**
We visualized results for prediction of the model using confusion matrix in Erosion (A) and JSN (B). The four points in Erosion and one point in JSN could not be displayed because there were very few teacher labels due to the constraints of SHS reading, which were difficult to train and the model could not regress and only a few were included in the test set.
Cor correlation coefficient, RMSE Root Mean Squared Error
CMC, carpometacarpal joint; PIP, proximal interphalangeal joint.

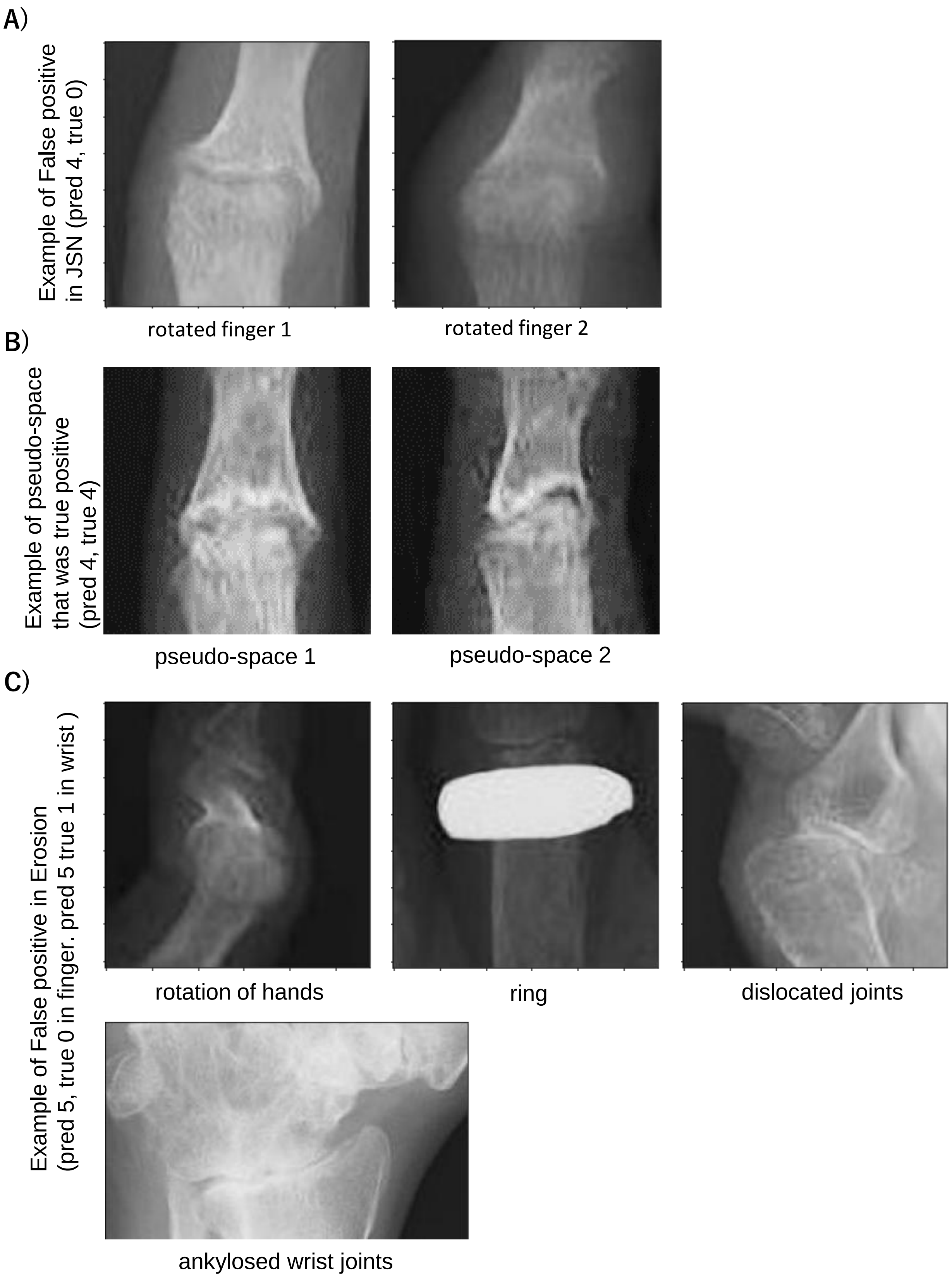


**Figure S8. Example of false positive with large differences in SHS scores.**
Our model mislabeled when the rotated joint was misclassified as pseudo-space (A). In the SHS method, a joint with a complete collapse that apparently retains a joint space is scored as pseudo-space with a JSN of 4 points. Our model could label pseudo-space as 4 points in JSN (B). Our model misjudged for rotated fingers, fingers with rings present, and some dislocated joints.(C). The scoring of the wrist joint with ankylosing is challenging task. Partial ankylosis of the wrist joint is scored 4 points in JSN, while it is scored depending on the degree of ankylosis in Erosion. In the example shown in the panel C, there is partial ankylosis, but the erosion score is smaller than it appears because the ankylosis does not extend to the entire wrist joint.

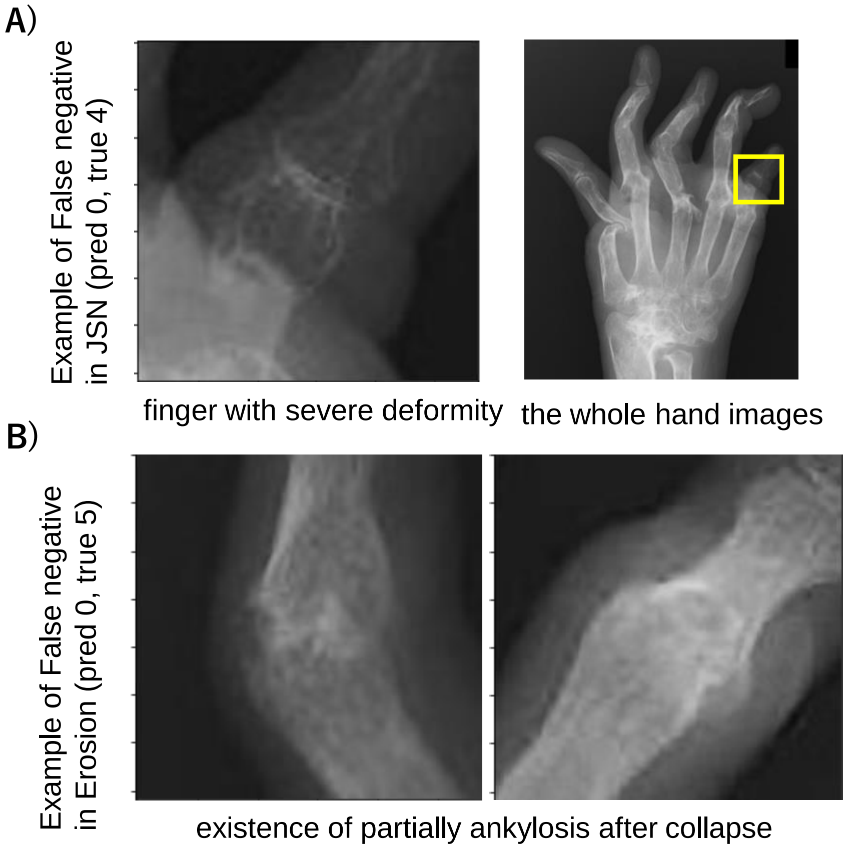


**Figure S9. Example of false negative with large differences in SHS scores.**
Our model misidentified joints with severe deformity and mislabeled it score 0 in JSN (A). The figure on the left is an enlarged view of the yellow square in the figure on the right. On the other hand, the model mislabeled joints with partially ankylosis and collapse score 0 in erosion (B).

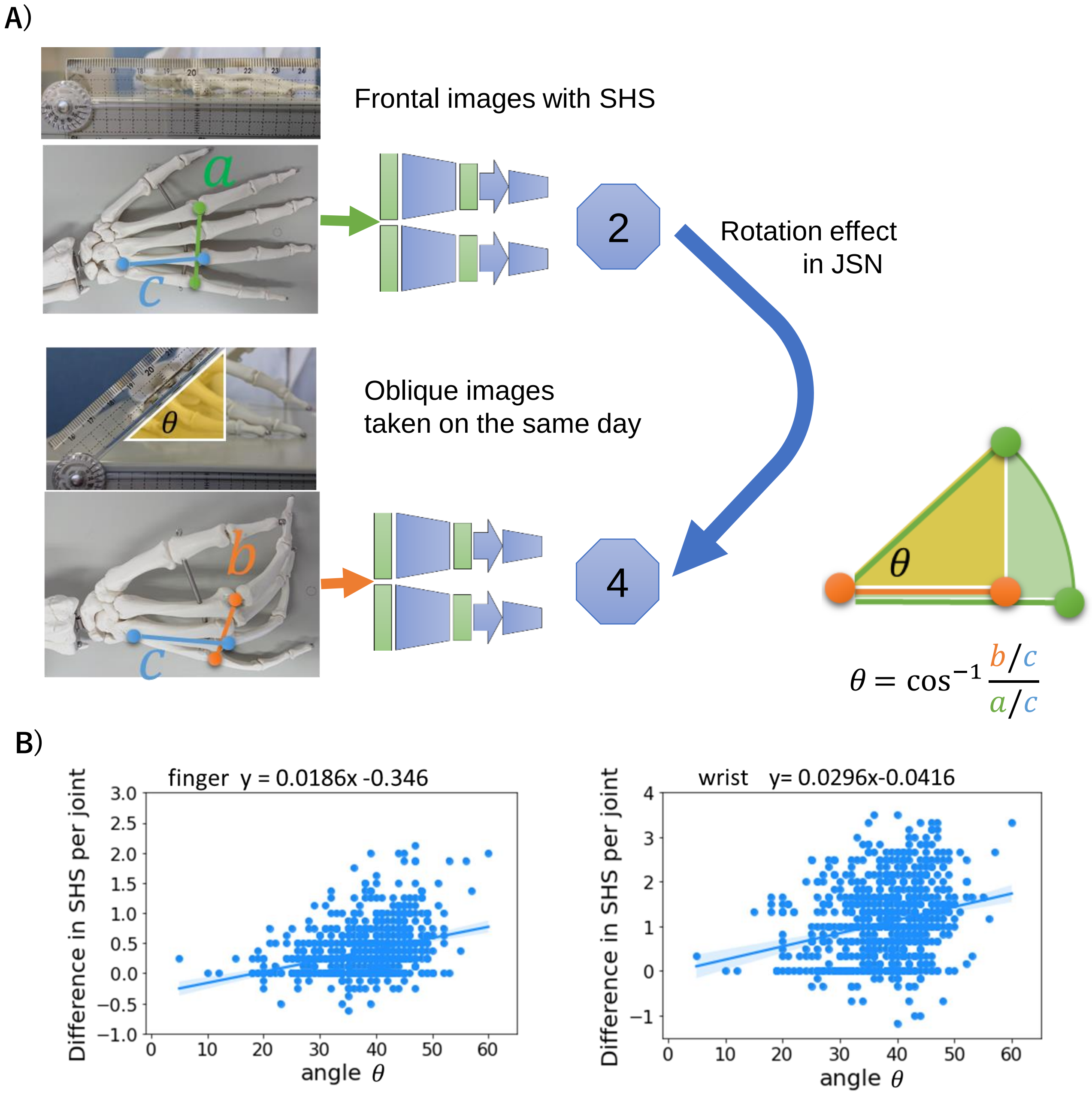


**Figure S10. Examination for effect of rotation effect on scores.**
We calculated the lengths of a, b, and c in Figure A based on the coordinates of the joints predicted by U-Net constructed in the Joint detection phase, and estimated the hand angle of the oblique image relative to the frontal image.(A). Results of the regression of the angle and the score difference per joint between the frontal and oblique images(B).

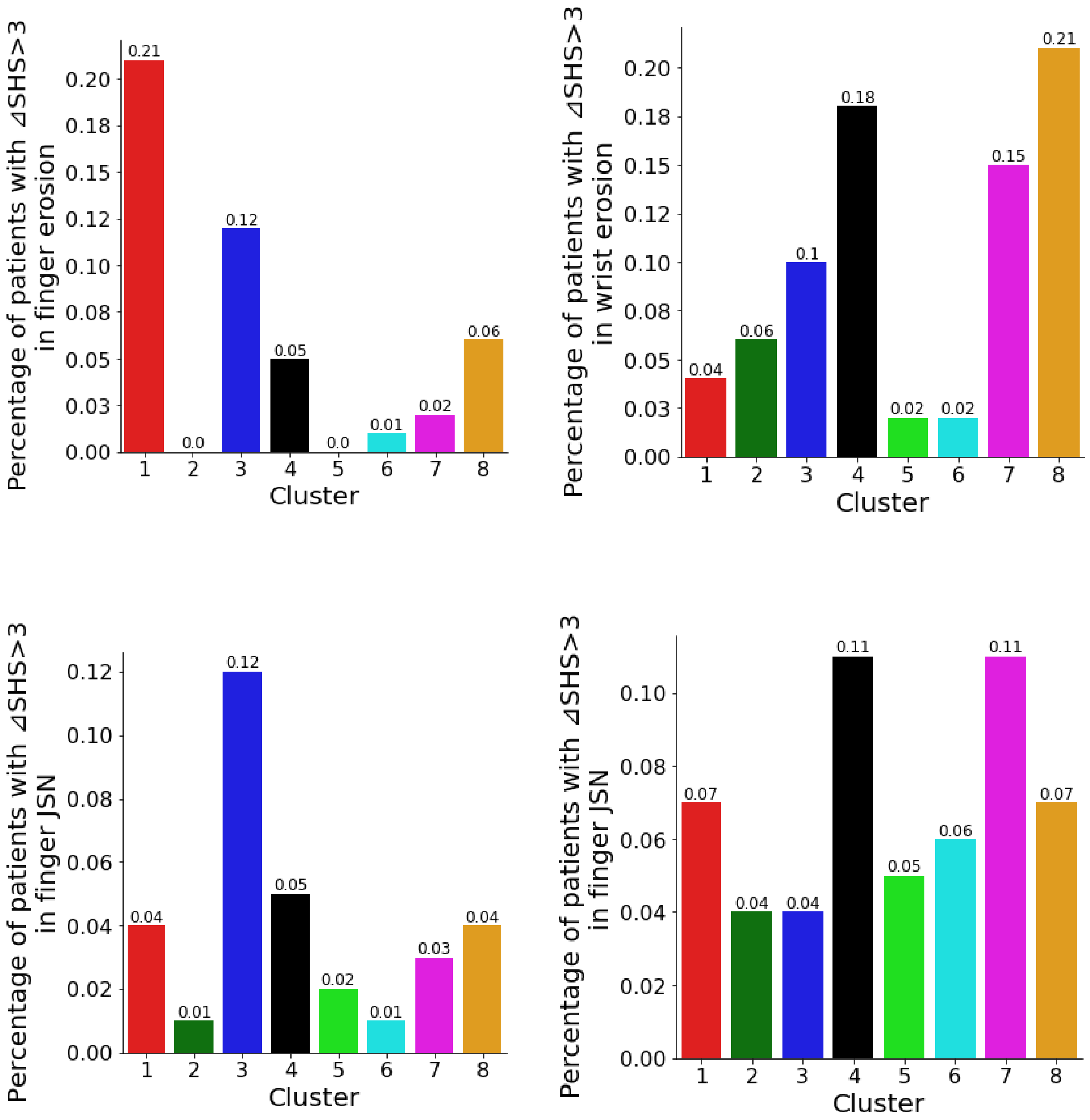


**Figure S11. Visualization of percentage of patients with progressive bone destruction.**
We defined ⊿SHS>3 as progression of joint destruction and showed the percentage of patients with progression in each cluster.
JSN, joint space narrowing.

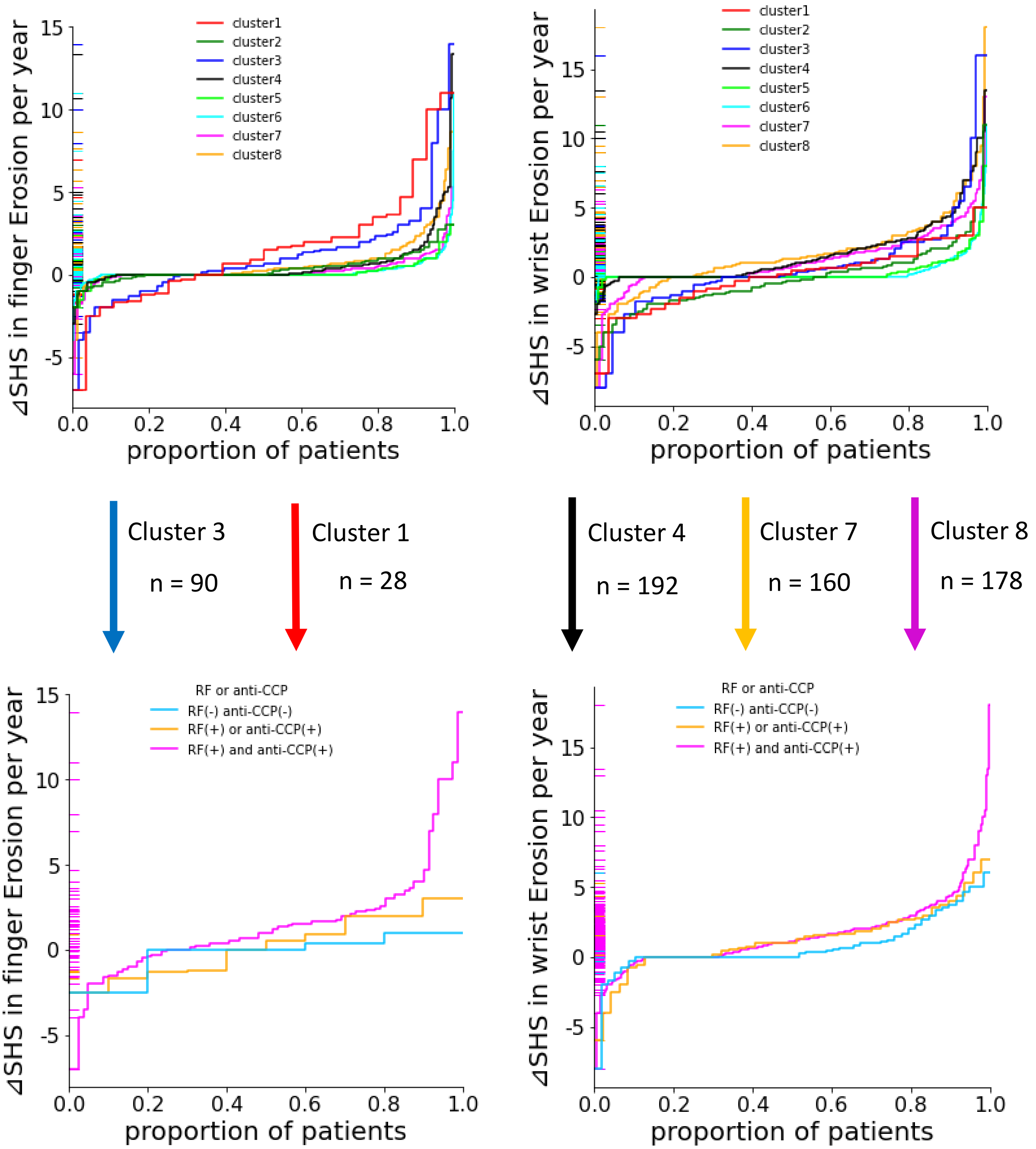


**Figure S12. Subgroup analysis stratified by serological marker after extraction of clusters**

We extracted clusters as shown in this figure and performed subgroup analyses according to serological markers.

CCP, cyclic citrullinated peptide; RF, rheumatoid factor; SHS, Sharp/ van der Heijde scores.
