## Supplementary table for "Development of a scoring model for the Sharp/van der Heijde score using convolutional neural networks and its clinical application in predicting radiographic progression using a graph convolutional network"

Supplementary table 1. Multivariable analysis for **∆**finger erosion

|  | coefficient § | standard error | 95% CI | | *P* value |
| --- | --- | --- | --- | --- | --- |
| Sex (female) | 0.0245 | 0.025 | -0.025 | 0.074 | 0.333 |
| Age onset | 0.0222 | 0.026 | -0.029 | 0.073 | 0.392 |
| anti-CCP or RF | 0.0688 | 0.026 | 0.019 | 0.119 | 0.007 |
| DAS28-CRP | 0.0992 | 0.026 | 0.048 | 0.151 | >0.001 |
| MTX (yes) | 0.0223 | 0.025 | -0.028 | 0.072 | 0.381 |
| csDMARDs (yes) ‡ | 0.0254 | 0.026 | -0.026 | 0.076 | 0.329 |
| bDMARDs (yes) | 0.0221 | 0.027 | -0.030 | 0.074 | 0.404 |
| Cluster 1 or 3* | 0.1853 | 0.026 | 0.134 | 0.237 | >0.001 |
| Cluster 4,7or 8† | 0.1096 | 0.026 | 0.058 | 0.161 | >0.001 |

* Patients with cluster 1 or 3 were coded 1 and remaining were 0.

† Patients with cluster 4,7 or 8 were coded 1 and remaining were 0.

‡ csDMARDs without MTX

§ Standardized regression coefficient

anti-CCP, anti-citrullinated peptide antibody; CI, confidence interval; DMARDs, Disease modifying anti rheumatic drugs; MTX, methotrexate; RF, rheumatoid factor.

Supplementary table 2. Multivariable analysis for **∆**wrist erosion

|  | coefficient § | standard error | 95% CI | | *P* value |
| --- | --- | --- | --- | --- | --- |
| Sex (female) | -0.0052 | 0.025 | -0.054 | 0.043 | 0.833 |
| Age onset | 0.0698 | 0.025 | 0.020 | 0.120 | 0.006 |
| anti-CCP or RF | 0.0497 | 0.025 | 0.001 | 0.099 | 0.048 |
| DAS28-CRP | 0.1095 | 0.026 | 0.059 | 0.160 | >0.001 |
| MTX (yes) | -0.0554 | 0.025 | -0.104 | -0.007 | 0.026 |
| csDMARDs (yes) ‡ | 0.0124 | 0.025 | -0.038 | 0.062 | 0.627 |
| bDMARDs (yes) | 0.0284 | 0.026 | -0.023 | 0.079 | 0.275 |
| Cluster1 or 3* | 0.0301 | 0.026 | -0.021 | 0.081 | 0.244 |
| Cluster4,7 or 8† | 0.2630 | 0.026 | 0.213 | 0.313 | >0.001 |

* Patients with cluster 1 or 3 were coded 1 and remaining were 0.

† Patients with cluster 4,7 or 8 were coded 1 and remaining were 0.

‡ csDMARDs without MTX

§ Standardized regression coefficient

anti-CCP, anti-citrullinated peptide antibody; CI, confidence interval; DMARDs, Disease modifying anti rheumatic drugs; MTX, methotrexate; RF, rheumatoid factor.

Supplementary table 3. Mean AUC from 10-fold-cross validation for each model and factor

|  | RF+ | anti-CCP | RandF | RandF_Boru | GCN |
| --- | --- | --- | --- | --- | --- |
| Mean AUC for wrist Erosion | 0.531±0.058 | 0.540±0.054 | 0.646±0.041 | 0.697±0.055 | 0.748±0.079 |
| Mean AUC for  Finger Erosion | 0.590±0.055 | 0.582±0.018 | 0.671±0.08 | 0.690±0.12 | 0.800±0.10 |

This table is presented in the form of AUC±SD (standard deviation)

anti-CCP, anti-citrullinated peptide antibody; AUC, area under the curve; Boru, Boruta; GCN, graph neural network; RandF, random forest; RF, rheumatoid factor.

Supplementary table 4. Top 10 ranked feature importance in prediction of ∆wrist erosion

| Rank | RandF | RandF_Boruta | GCN |
| --- | --- | --- | --- |
| 1 | L_J_capitate | cluster4,7,8 | RF |
| 2 | L_J_trapezium | L_J_capitate | ESR |
| 3 | L_J_radius | R_J_ trapezium | anti-CCP |
| 4 | CRP | L_J_trapezium | L_J_trapezium |
| 5 | R_J_radius | SDAI | age_onset |
| 6 | cluster4,7,8 | R_J_capitate | R_J_capitate |
| 7 | SDAI | duration | L_J_PIP4 |
| 8 | duration | R_J_radius | CRP |
| 9 | R_J_capitate | R_E_radius | cluster4,7,8 |
| 10 | anti-CCP | ESR | L_E_PIP4 |

anti-CCP, anti-citrullinated peptide antibody; CRP, C-reactive protein; DAS, Disease Activity Score; E, erosion; ESR, erythrocyte sedimentation rate; J, joint space narrowing; L, left; PIP, proximal interphalangeal joint; R, right; RandF, random forest; RF, rheumatoid factor; SDAI, Simplified Disease Activity Index.

Supplementary table 5. Top 10 ranked feature importance in prediction of ∆finger erosion

| Rank | RandF | RandF_Boruta | GCN |
| --- | --- | --- | --- |
| 1 | DAS28ESR | DAS28ESR | anti-CCP |
| 2 | duration | anti-CCP | RF |
| 3 | anti-CCP | ESR | Age onset |
| 4 | ESR | R and L ulna_d | ESR |
| 5 | SDAI | duration | CRP |
| 6 | R_J_capitate | SDAI | R_J_MCP1 |
| 7 | R and L ulna_d | R_J_MCP1 | R_J_MCP2 |
| 8 | DAS28CRP | DAS28CRP | L_J_MCP2 |
| 9 | R_ulna_d | L_ulna_d | R_J_PIP3 |
| 10 | R_J_MCP1 | R_J_capitate | L_J_PIP5 |

anti-CCP, anti-citrullinated peptide antibody; CM, carpometacarpal joint; CRP, C-reactive protein; DAS, Disease Activity Score; E, erosion; ESR, erythrocyte sedimentation rate; J, joint space narrowing; L, left; MCP, metacarpophalangeal joint. PIP, proximal interphalangeal joint; R, right; RandF, random forest; RF, rheumatoid factor; SDAI, Simplified Disease Activity Index; ulna_d, ulnar deviation.
