## Supplementary methods for "Development of a scoring model for the Sharp/van der Heijde score using convolutional neural networks and its clinical application in predicting radiographic progression using a graph convolutional network"

Contents:

Supplementary methods

**Supplementary methods**

**Study population.** All patients were enrolled in the Institute of Rheumatology, Rheumatoid Arthritis (IORRA) cohort and clinical information was obtained from this cohort. IORRA cohort is a single, institute-based, large, observational cohort of Japanese patients with RA, and includes about 1% of all Japanese RA patients. All radiographs were extracted from IORRA Navigation Database of RA image (INDRA) which is the largest database in Japan linked to clinical data, consisting of approximately 50,000 radiological images of 66 different types taken between 1997 and 2021.

**Evaluation of radiographic joint damage scored by specialist.**

We collected the data for 1,342 patients of the Sharp/van der Heijde score (SHS) [1] from the medical records of patients. SHS, which was scored by a single experienced evaluator (K.Y) with proper anteroposterior radiographs of the hands, was used to evaluate radiographic joint damage. In section for verification of applicability, we calculated differential SHS for a patient using the following formula,

$$\Delta SHS = (SHS in year 2 - SHS in year 1)/(year 2 - year 1)$$

, where $year 1$represents the earliest year of the radiographic date of a given patient within INDRA, and $year 2$ represents the closest radiographic date to five years after $year 1$. In section for construction of prediction model, we defined the radiographic progression group as $\Delta SHS$ > 3 and the non- radiographic progression group as the remaining patients.

**Preparation for heatmap regression.**

We generated heatmap images which encode the “pseudo-probability” of joint coordinate being at a particular pixel location. This method is also used for cell segmentation [2], and we referred to it as well. To generate heat map images from the coordinates, we used a Gaussian filter, which is a type of smoothing filter that performs smoothing by blending surrounding pixels. The Gaussian filter uses a Gaussian function for the weights to reduce the effect of smoothing on pixels that are distant from the reference pixel. The weight by the Gaussian function in generating the 2D heat map image is defined as the Gaussian function *G_i_ (x, y,σ)* with the coordinate value (x, y) of a certain pixel as follows.

*G (x, y,σ)* =$\frac{1}{2\pi\sigma^{2}}exp\left( -\frac{\left( x^{2}+y^{2} \right)^{2}}{{2\sigma}^{2}} \right)$

the standard deviationσis a parameter that controls the width of the Gaussian function. Too wide a value of σ may cause deviations in the prediction of landmark locations, while too small a value may impair the learning process and prevent the generation of predictive heat maps.

The region where this Gaussian weighting is applied is then called the kernel, and the Gaussian filter is defined as follows.

*g (i, j)* = $\frac{1}{W}\sum_{n=-k}^{k} \sum_{m=-k}^{k} G\left( n,m,\sigma\right)f(i+n,j+m)$

*W =* $\sum_{s=-k}^{k} \sum_{t=-k}^{k} G\left( s,t,\sigma\right)$

where k is half the kernel size (e.g. if the kernel size is 101, then k = 50), g(i,j) is the pseudo-probability value of the output coordinate (i, j), and f(i+n, j+m) is the value of the coordinate (i+n, j+m), which is n pixels distant to the x-axis and m pixels distant to the y-axis from the output coordinate. The surrounding pixels (i+n, j+m) are weighted and added together based on a Gaussian function. To prevent the sum from increasing excessively, divide the sum of the weighted pixels by the weights. Based on the results of our preliminary experiments, we generated heatmap images from coordinates with kernel size 201 and sigma 6 by using Python OpenCV Gaussian Blur filter. Since 15 landmarks are set for a single hand radiograph, 15 correct heat map images are generated for a single training image.

**Convolutional neural networks.**

*Orientation phase.* We adopted pre-trained Efficient net b0 [3], a lightweight, high-performance model, because it provided 100% accuracy (Extended figure S1). We trained the model with the images resized to 144 × 144 pixels for both the one and two hand images. We used Adabound [4] as the optimization algorithm and used Cosine-Annealing [5] as learning scheduler (lr = 0.01-0.0001, T_max = 5) for 10 epochs. The loss function was set to softmax cross-entropy, and the batch size for the training was set to 16.

*Joint detection phase.* We initially built models that directly regressed coordinates using ResNet [6], DenseNet [7], or EfficientNet, but they did not reach an accuracy that could be adapted to the Damage prediction phase. Therefore, we performed weakly supervised learning by heat-map regression based on U-Net [8]. We generated heat maps as described above (see Preparation for heatmap regression in Methods and Extended figure S4) from the correct labels of the joint coordinates for each image. We first trained a standard U-Net model (Extended figure S3) with the following hyperparameters (optimizer: Adam, scheduler: Cosine-Annealing (lr = 1.0e-03-1.0e-06, T_max = 1000), loss function: mean squared error, epoch: 1000, batch size: 16, Augmentation: random rotate, flip and changes from hue, saturation, brightness and contrast) and achieved best score. We incorporated squeeze-and-excitation blocks and coordconv into the standard U-Net and changed the loss function from Mean Squared Error (MSE) to Wing Loss[9], but the performance of the model was almost the same or even decreased compared to the standard U-Net model (data not shown).

*Damage prediction phase.* We tested several CNN models (VGG [10], ResNet, DenseNet, Xception [11], EfficientNet), and adopted EfficientNet because it provided the highest performance (data not shown). We then constructed full model combined with various modules follow; Over-sampling [12], Transfer learning, squeeze-and-excitation blocks[13], self-mutual learning[14], Adabound and Cosine annealing and compared full model with models removed or replaced modules from full model (figure3B and Extended figure S6). To compare all models on the same test images, we first divided each joint image dataset into 10 segments, used one as a test set, and cross-validated with the rest. We ensembled the nine models constructed in cross validation and predicted the test set. The model “downsampR_0.4” and “downsampR_0.2” mean that 40% and 20% of the original images were used for training, respectively. Note that in this case, after making one-tenth of each joint image dataset as the test-set, we randomly extracted 40% or 20% images for cross-validation. For oversampling, after creating a test set of one-tenth of each joint image data set, we oversampled the remaining images so that the number of non-score 0 images was equal to 20% of the number of score 0 images. For example, if there were 100 images with a score of 0, 5 images with a score of 1, and 10 images with a score of 2, after Oversampling, there would be 20 images for both score 1 and score 2. The reason for the 20% oversampling was that increasing the percentage further would have reduced the performance. Unlike the wrist joints, model with oversampling had lower performance for predicting finger erosion, PIP and MCP in JSN. The number of image data for the wrist joint differs from that for the finger joint by a factor of up to 10, which may be caused by the obvious difference in data structure. For this reason, Oversampling was not used for predicting finger erosion, PIP and MCP in JSN.

We trained a full model EfficientNet with the following hyperparameters (optimizer: Adabound, scheduler: Cosine Annealing with T-max = 40 and learning rate from 5e-03 to 1e-06, loss function: cross entropy, epoch: 200, batch size: 32, Augmentation: RandomRotation, ColorJitter (brightness=0.5, contrast=0.5, saturation=0.5, hue=0.5), RandomPerspective). For finger damage prediction, only one score is predicted from a single joint image, so the model has only one fully connected layer. On the other hand, for wrist damage prediction, a multi-task model was employed to predict multiple joint scores from a single wrist image. That is, in the case of predicting the wrist narrowing score, the model has three fully connected layers (FCL) to predict three scores from a single wrist image (3CM-4CM-5CM or Capitate-Trapezium-Radius), while in the wrist erosion score, the model has two FCL to predict two scores from an image (Metacarpal-Trapezium, Lunate-Scaphoid, or Ulna-Radius).

**Assessment for image rotation effect**

To test the rotation effect on scores, we estimated the angle formed by the straight line connecting the 2MCP and 5MCP in the frontal and oblique images as follow (Extended figure S10). First, we extracted the frontal image with SHS and the oblique image taken on the same day from INDRA. Next, we calculated angle using following formula,

$$\theta=\cos^{-1} \frac{b/c_{\mathrm{oblique}}}{a/c_{\mathrm{frontal}}}$$

,where $a$ is the length of the line from 2MCP to 5MCP in the frontal image, $b$ is the length of the line from 2MCP to 5MCP in the oblique image, $c_{\mathrm{frontal}}$is the length of the fourth metacarpal bone in the frontal image $c_{\mathrm{oblique}}$ is the length of the fourth metacarpal bone in the oblique image. Due to the difference in image size between frontal and oblique, a and b cannot be substituted into the same equation, so c is used for normalization of a and b.

**Graph Convolutional Network**

Our GCN model was built on previous model[15,16]. Our graph network considers each patient as a node in a network connected by edges based on cosine similarity. Here is a formal definition of the graph

$G=(V,E)$	(1)

, where $V$ represents the vertex set consisting of nodes $\left\{ v1,. . . ,vn \right\}$, and $E$ represents an edge based on neighborhood relationship. This graph $G$ can be described using an adjacency matrix $A(\in\mathbb{R}^{n\times n})$. If $A(i, j)$ is 0 indicates that there is no edge between node $vi$ and node $vj$, otherwise there is an edge between the two nodes and the value means weight of the edge. In our model, each node represents a patient and has d-dimensional feature vectors (ex. $f1$: age, $f2$: sex...$fd$: CRP). Thus, the node feature matrix is denoted by $X\in\mathbb{R}^{n\times d}$. A and X are substituted into the function of the GCN model where each layer is defined as follows

$H^{(l+1)}=f(H^{(l)},A)=\sigma(AH^{(l)}W^{(l)})$	(2)

, where $H^{(l)}$ is the input of the lth layer and $H^{(0)}$is the input of layer 0, thus $H^{(0)}$=$X$. $W^{(l)}$ $\left( \in\mathbb{R}^{s\times s+1} \right)$ is the trainable weight matrix of layer l, and σ represents an activation function and Leaky ReLU was employed in this study. Before substituting the adjacency matrix A into the function of GCN model (2), it is converted to a Laplacian matrix shown below,

$\mathcal{L}=\hat{D}^{-\frac{1}{2}}\tilde{A}\hat{D}^{-\frac{1}{2}}$	(3)

Here, 𝐴̃ = 𝐴 + 𝐼 is the sum of the adjacency matrix and the identity matrix. Since the diagonal elements of the adjacency matrix are all zeros, the result of the inner product of this adjacency matrix $A$ and $H^{(l)}$ will capture the information of neighbor nodes but lose the information of own nodes. By adding $I$ to $A$, the feature vector of each node can be updated while retaining its own information. $\hat{D}$ is the degree matrix of the 𝐴̃. The Laplacian has the role of smoothing their features because the feature size of densely connected nodes increases.

Current GCNs are known to lose accuracy as they become multi-layered due to over-smoothing issues. In our preliminary experiments, the GCN consisting of two layers also had the highest accuracy. Thus, our GCN consists of an input layer ($l$=0) and an output layer ($l$=1). We inserted a dropout layer between these two GCN layers, and finally placed fully connected layers with a Leaky ReLU (figure 7). The number of dimensions of the weight matrix was set to be reduced by half at each GCN layer, i.e., $W^{(0)}\in\mathbb{R}^{d\times\frac{d}{2}}$ and $W^{(1)}\in\mathbb{R}^{\frac{d}{2}\times\frac{d}{4}}$. We adopted cross-entropy loss as loss function.

In our GCN model, nodes are connected with edges when the cosine similarity between them is larger than a threshold$\epsilon$. $Aij$, the adjacency between $vi$ with feature vector $xi$and $vj$with feature vector $xj$ in the graph, is calculated as follows,

$Aij=\left\{ \begin{aligned} \frac{xi\cdot xj}{\left\| xi \right\|_{2}\left\| xj \right\|_{2}} \\ 0, otherwise \end{aligned} \right., if i\neq j and \frac{xi\cdot xj}{\left\| xi \right\|_{2}\left\| xj \right\|_{2}}\geq\epsilon$	(4)

The threshold $\epsilon$ is defined as the $k\times nth$ cosign similarity among the $n\times n$ sorted cosine similarities. $k$ is a hyperparameter and represents the average number of edges per node. Therefore, the case $k$= 1 constitutes a graph with no edges between nodes because all the top 1 × $n$ cosine similarities are values due to self-connecting.

When testing new samples, we extended the graph consisting of trained samples by adding validation samples to the graph as previously reported [15]. First, the GCN network is trained according to the following equation,

$\hat{Y}_{tr}=GCN(X_{tr}, \tilde{A}_{tr})$	(5)

, where $\hat{Y}_{tr}\in\mathbb{R}^{n_{tr}\times2}$ represents the predicted label probability of the training sample for classification of the two classes (patients with and without progressive erosion). When we predict the radiographic progression for validation sample of $n_{val}$, we extended the node feature matrix to $X_{tr+val}\in\mathbb{R}^{{(n}_{tr}+n_{val})\times d}$ and adjacency matrix to $\tilde{A}_{tr+val}\in\mathbb{R}^{{(n}_{tr}+n_{val})\times{(n}_{tr}+n_{val})}$ and assigned to GCN network as follow,

$\hat{Y}_{tr+val}=GCN(X_{tr+val}, \tilde{A}_{tr+val})$	(6)

, where $\hat{Y}_{tr+val}\in\mathbb{R}^{{(n}_{tr}+n_{val})\times2}$ represents the predicted label probability of the training and validation sample. The weights in training were fixed and used in validation. Since the hyperparameter k is also the same for training and validation, the structure of nodes and edges in the training graph remains unchanged, and edges and nodes of new validation sample are added to the training graph.

We trained our GCN model with the following parameters, Leaky ReLU slope of 0.1 and drop rate of 0.6.

**Calculation of feature importance in GCN**

We employed the *Ablation method* as in previous reports [15]. Briefly, we removed the signal from that feature by setting the vector of a feature to 0 and considered the feature resulting in a lower AUC as important. Since we employed a 10-fold cross validation to evaluate the model, we calculated the feature importance by summing the AUC decrease at each fold. Note that the calculation method of feature importance for GCN is different from those for RF and cannot be compared.

**Random Forest**

To compare the performance of GCN, we used the Random Forest (RF). We adjusted each set of hyperparameters using k-fold cross-validation with GridSearchCV module from scikit-learn as follow, max depth (3,4,5), min samples split (3,4), max leaf nodes (4,5), max features (log2, sqrt), and n estimators (20, 60, 100, 140, 180).

For Random Forest, we also evaluated models applying Boruta, a feature selection method [17]. Briefly, Boruta create fake features that should not contribute to performance of classification. By comparing the feature importance of each feature with that of the fake features, we can select more important features for classification after training RF including the fake features. We set hyperparameters of Boruta, max_iteration and perc, to 800 and 70, respectively.

**Statistical analysis.**

Missing values were imputed using missForest [18], a random forest imputation method. We conducted hierarchical clustering using the Seaborn Clustermap function with ward as the method and euclidean as the metric. A multiple regression model was used for multivariable analysis. Quantitative variables were normalized before applying the model. Clusters were binarized as follow, Cluster 1 or 3: patients with cluster 1 or 3 were coded 1 and remaining were 0, Cluster4, 7 or 8: patients with cluster 4,7 or 8 were coded 1 and remaining were 0. Variance inflation factor (VIF) was used to check for multicollinearity among the covariates using Python. We performed 10-fold cross-validation (training : testing ratio, 9 : 1) to evaluate the performance of Random Forest with and without Boruta, and GCN model. We used pingouin (ver 0.5.1, open-source statistical package written in Python 3) to calculate ICC3 of ∆SHS between models and experts reading.

**Environment.**

All the DL models were created and trained using pytorch libraries (version 1.7.1) in a Python 3.8.12 environment. All the trainings and further analysis were performed on a single workstation with Nvidia GeForce RTX 3090 (24Gb)
